# TSGMA: identification of macro associations from global data to build global MA networks

**DOI:** 10.1101/2025.06.09.25329257

**Authors:** Hongyue Ma

## Abstract

The growing availability of globally harmonized datasets offers unprecedented opportunities to identify population-level risk factors, yet systematic tools for macro-scale association analysis remain scarce. Here, I introduce the concept of “macro association” (MA) and propose a three-tiered framework: Three Stars Global Macro Association-analysis (TSGMA), which integrates correlation, partial correlation adjusted for confounders, and temporal lag analysis to rapidly and robustly rank global associations. Applying TSGMA to dietary and health data from 159 countries or regions, I identify strong links between diet and three cardiometabolic markers. Animal fats group, red meat, and eggs show robust, time-lagged associations with elevated non-HDL cholesterol; sugar, cereals, and poultry meat are associated with increased diabetes prevalence; while starchy roots and pulses consistently exhibit protective associations. TSGMA not only confirms established patterns but also reveals overlooked signals, offering a scalable approach to construct integrative global “MA networks” and enabling hypothesis generation from open-access “macro data”.

**Graphical Abstract:** 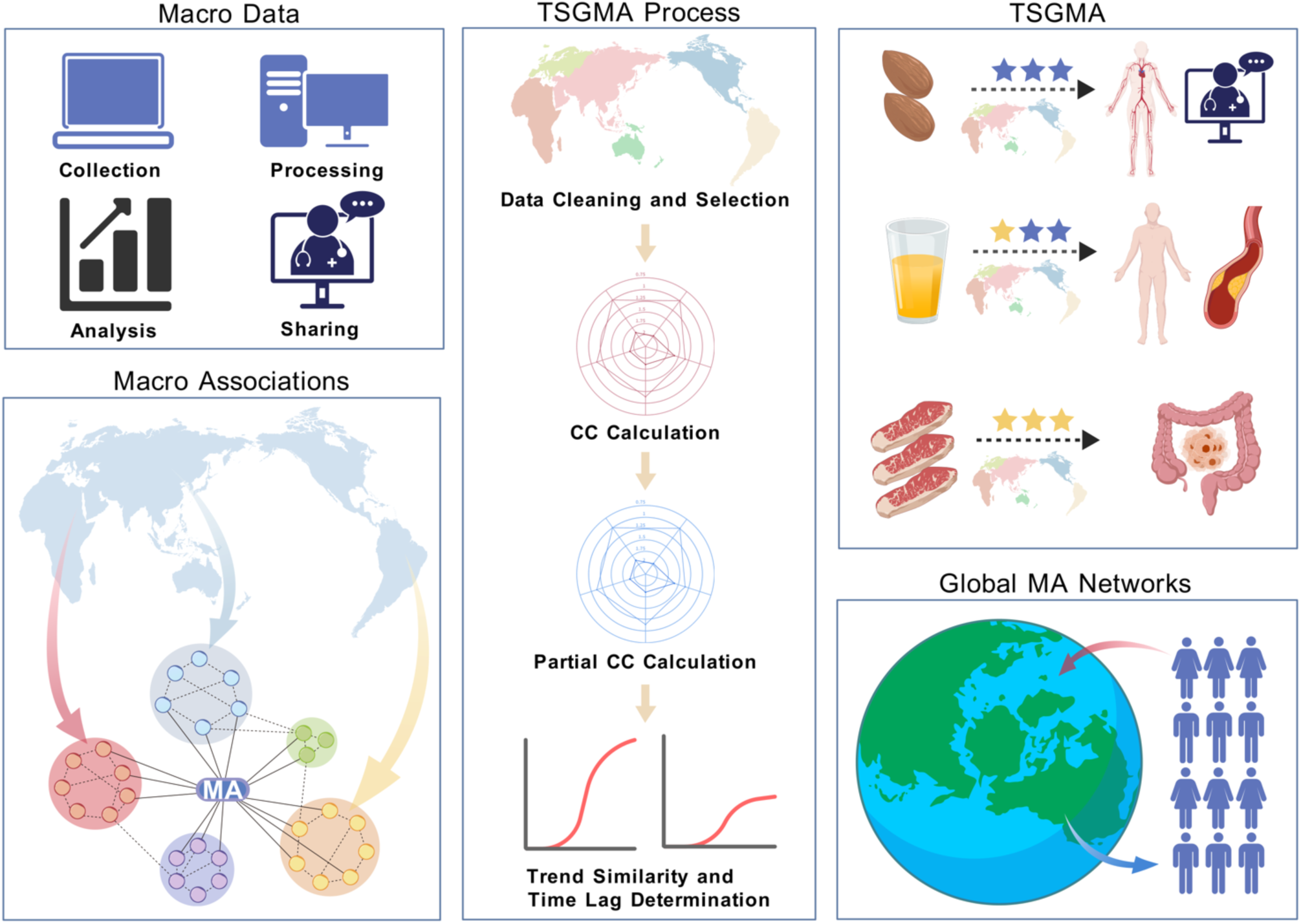

## 1. Introduction

Understanding the myriad factors that influence human health is a central challenge in science^1^. Lifestyle and environmental factors such as diet, medications, physical activity, sleep, air quality, and work conditions interact in complex ways to affect health outcomes^1–3^. Broadly mapping these relationships can greatly improve our ability to prevent disease and promote wellness. Identifying potential risk factors and protective factors through comprehensive association analysis can guide more focused research and interventions.

Historically, early epidemiology relied on simple ecological correlations or rule-of-thumb observations^4^. The most famous classic example is the discovery of geographic consistency between smoking prevalence and lung cancer mortality in the middle ages in the twentieth century, which proved the heuristic value of crude analyses^5^. But since then, limitations resulting from issues such as sparse data, rudimentary statistics, and profound confounding restricted both scope and credibility have come to light^6–8^. Over the following decades, controlled trials, large prospective cohorts, and meta-analyses transformed risk assessment into a rigorous discipline^9–11^. Landmark resources such as the UK Biobank and the Global Burden of Disease study now permit precise causal inferences to be made about prespecified hypotheses^1,10^. However, this depth comes at the cost of breadth: cohorts are expensive to assemble, most cover a narrow range of populations, and by design they can only test a limited group of candidate exposures^10,12–14^. As a result, modern studies are better at confirming known risks but less adept at identifying new ones.

Meanwhile, digitization has generated an unprecedented number of open national statistical databases, such as Our World in Data (https://ourworldindata.org), the World Bank (https://data.worldbank.org), FAO (https://www.fao.org/faostat/en/#data/FBS) and International Agency for Research on Cancer (https://www.iarc.who.int), as well as unprecedented conditions for data collection. These “macro data” (MD)are numerically simple, globally harmonized and directly interpretable by both scientists and the public, however, they are still lack being systematically mined^15–17^. Traditional fine-grained epidemiological methods are not applicable to the analysis of these data, and exploratory, simple correlation analyses have been gradually abandoned as yielding results that are inferior to those of precise epidemiological studies^6,8,15,17–19^. Broad exploratory studies of cross-cutting risk factors, once central to public health research, are increasingly seen as too “ecological” or methodologically difficult to navigate^19–21^. Unfortunately, it is in the very era that provides the richest data that comprehensive, integrated broad explorations of risk factors are retreating.

Thus, against the backdrop of data richness that exceeds any previous era, I propose a novel concept: “macro association” (MA), which I define as a wide range of repeatable associations detectable on a domain-wide scale in the context of MD. Moreover, I name the systematic analytical mining of MA as “macro association analysis” (MA-analysis), hence, MA-analysis aims to uncover such associations in large publicly available datasets as well as in various accessible data sources, and ultimately to build a comprehensive domain-wide “macro association network” (MA network) across various domains, such as health, economy, environment, climate, and other relevant domains. Among the numerous possible dimensions of MA, the country-based global dimension is currently the most attractive due to the increasing completeness of the data, the simplicity of the figures, and the direct relevance to human policies. However, simple correlations are easily distorted by confounding and reverse causation, and gold standard epidemiological methods are not applicable to the analysis of this type of data, making MA-analysis in these data particularly difficult^6,22^. To bridge this gap, I developed the Three Stars Global Macro Association-analysis (TSGMA), a three-step screening methodology that grades pairs of associations between any global variables from zero to three stars by sequentially applying crude correlations (i, one star), confounder-controlled correlations (ii, two stars), and time lag analyses (iii, three stars). TSGMA provides an accurate, fast, and efficient way to perform MA-analysis of global dimensional data on a country basis, and lays the foundation for the construction of global macro association networks.

As a proof of concept, I applied TSGMA to the analysis of associations between dietary components and three cardiometabolic markers (hypertension, diabetes, and non-HDL-C levels) in 159 countries or regions (1992-2017). The methodology recovered typical associations, such as those between sugar and diabetes as well as alcohol and hypertension. At the same time, it revealed some overlooked patterns, such as a population lag of about 15 years between animal fat intake and non-HDL-C levels. These results illustrate how TSGMA can translate global MD into actionable hypotheses and lay the groundwork for building comprehensive, cross-cutting global MA networks.

In addition, I propose a structured framework for the construction of a global MA network. The process begins with applying TSGMA to individual databases to build internal MA networks, which are then progressively extended across multiple repositories. These independent networks are subsequently harmonized and iteratively integrated using artificial intelligence and advanced analytics into a unified global MA network. This framework transforms decentralized, open-access datasets into a cohesive, dynamic system capable of uncovering the intertwined drivers of planetary health, development, and sustainability.

## 2. Methods

### 2.1 Overview of the TSGMA approach

TSGMA is a three steps analytical methodology designed to explore and categorize associations between variables accurately and quickly with global data such as multiple years of country or region level data. Figure 1 provides a schematic overview of the TSGMA workflow. Briefly, the steps are as follows: (1) One star analysis (correlation analysis): Correlation coefficients between variables are calculated to screen for significantly correlated variables (*p* < 0.05). (2) Two stars analysis (partial correlation analysis): Key confounding variables are adjusted using partial correlation analysis to assess whether the observed associations persist after controlling for confounding effects (*p* < 0.05). (3) Three stars analysis (temporal correlation analysis): Temporal patterns of paired variables are examined to determine whether the change in one of the variables is consistently similar to the trend of the other variable with a relatively stable time lag. At each step, candidate variables are screened and given a “score” to indicate the strength of evidence for a true association: “zero star” = no association, “one star” = suggestive association, “two stars” = robust association after confounder adjustment, and “three stars” = probable causality with temporal priority.

**Figure 1.**
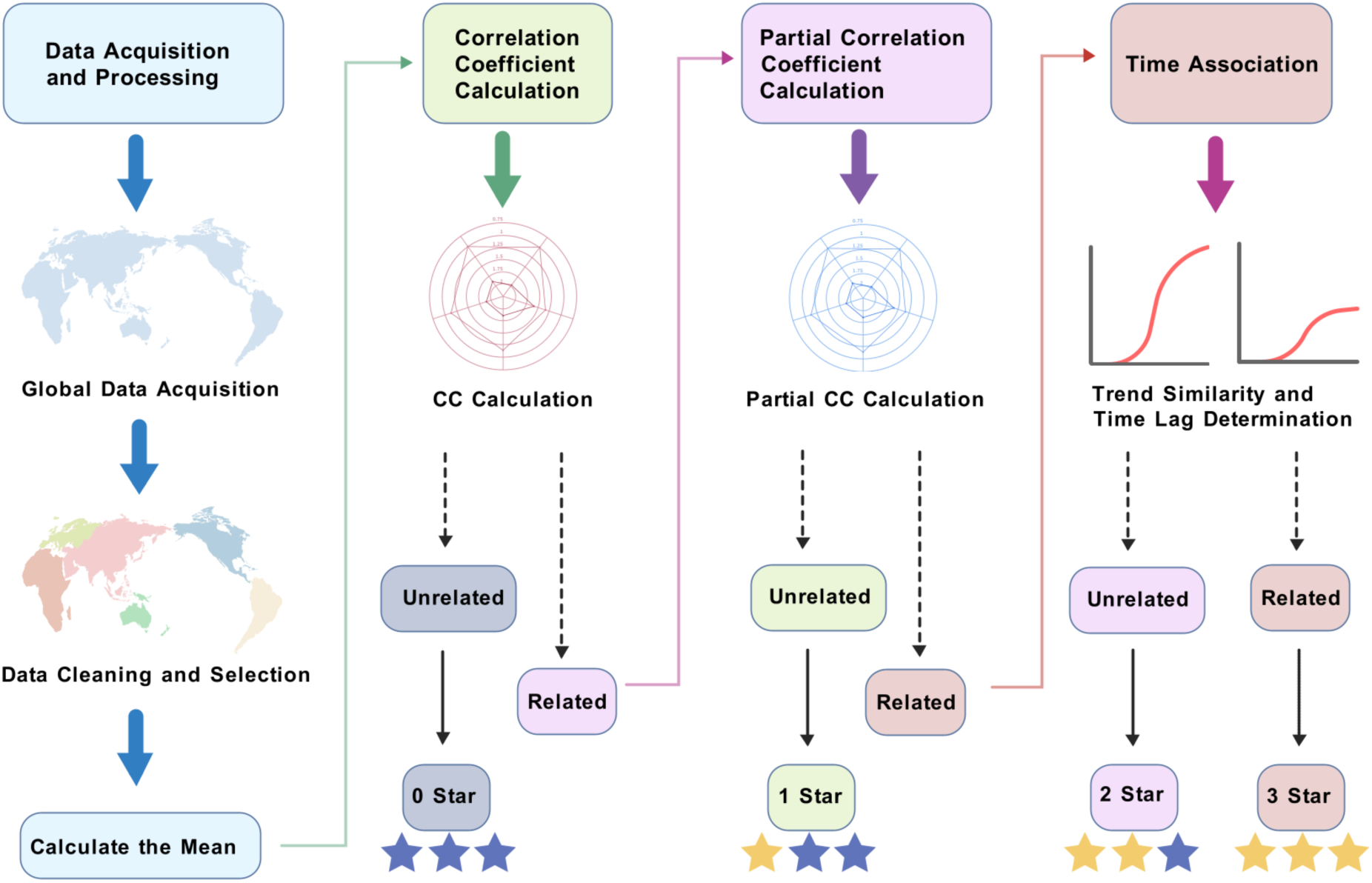
Overview of the TSGMA workflow. (i) Define the scope of the analysis and identify all variables of interest. (ii) Collect data for each variable from all countries or regions for which data are available. Data are cleaned and screened for countries or regions whose data are as complete as possible for a given period, and the data for each variable are then averaged. (iii) Calculate the correlation coefficient between each candidate variable. Pairs of variables showing significant correlation (*p* < 0.05) are screened for two stars analysis and uncorrelated variables (zero star) are discarded. (iv) Identify key potential confounders or key confounder representations based on the properties and meaning of the variables (for global variables, a classic example is socio-economic status, where GDP per capita is a useful key representation indicator^23^). Partial correlation coefficients between one star associated variables are calculated to control for confounders. Variables that maintain significant associations (*p* < 0.05) are upgraded to two stars associations, while those that lose significance are considered only at the one star association level. (v) A time-lag analysis is conducted for each two stars association: comparing the time trends of the two variables across multiple countries or regions on a country or regional basis. If the trends of the two variables are similar in a significant number of countries or regions and there is a relatively stable time lag relationship between the two, this indicates a potential causal relationship, which is categorized as a three stars association. If no clear temporal relationship is observed, the association remains at the two stars level.

### 2.2 Data source and selection

Dietary composition data in calories for 193 countries or regions for 1961-2022 from Our World in Data (https://ourworldindata.org/grapher/dietary-composition-by-country?country=~OWID_WRL) database was obtained, which were categorized into Alcoholic beverages, animal fats group, vegetable oils, oil crops, fish and seafood, sugar crops, sugar and sweeteners, starchy roots, other meat, sheep and goat, pig meat, poultry meat, beef, eggs, milk, nuts, fruit, vegetables, pulses, barley, maize, rice, wheat, other cereals, miscellaneous group (Share of dietary energy supplied by food commodity types in the average individual’s diet in a given country or region, measured in kilocalories per person per day). Due to missing data for some countries or regions, data for 159 countries or regions from 1992-2017 were selected in order to cover as many countries or regions as possible and over a longer period of time. All major dietary categories were further categorized into the main 16 groups: red meat, alcoholic beverages, animal fats group, milk, eggs, sugar and sweeteners, vegetable oils, nuts, poultry meat, vegetables, pulses, cereals, fish and seafood, starchy roots, fruit and other meat, of which barley, maize, rice, wheat, other cereals comprise “cereals”, pig meat, beef, sheep and goat comprise “red meat”. Other dietary components were not counted due to minimal intake and not being the mainstream dietary component. Specific information on these 16 dietary components is included in Table 1.

**Table 1.** Dietary components details.

| Dietary components | Food items |
| --- | --- |
| Red meat | Pork, beef, sheep and goat meat |
| Alcoholic Beverages | Beer, wine, spirits and other alcoholic beverages |
| Animal fats group | Lard, Tallow, Butter, Ghee, Schmaltz, Duck Fat, and Bacon Fat |
| Milk | dairy products made from milk ingredients, except butter |
| Eggs | Eggs |
| Sugar and Sweeteners | Brown Sugar, Powdered Sugar, Raw Sugar, Honey, Maple Syrup, Agave Syrup, Corn Syrup, Stevia and Artificial Sweeteners |
| Vegetable Oils | Soybean Oil, Canola Oil, Olive Oil, Sunflower Oil, Corn Oil, Palm Oil, Coconut Oil, Peanut Oil, Sesame Oil, Grapeseed Oil, Rice Bran Oil |
| Nuts | Almonds, Walnuts, Cashews, Pecans, Hazelnuts, Pistachios, Macadamia Nuts, Brazil Nuts, Pine Nuts, and Peanuts |
| Poultry meat | Chicken, Turkey, Duck, Goose, Quail, and Guinea fowl |
| Vegetables | Carrots, Beets, Spinach, Broccoli, Onions, Tomatoes, Bell Peppers, Cucumbers, Garlic, Zucchini, Eggplant, etc. |
| Pulses | Lentils, chickpeas, peas, kidney beans, black beans, pinto beans, navy beans, black-eyed peas, mung beans, cannellini beans. |
| Cereals | wheat, barely, rice, maize, others |
| Fish and seafood | all fish species and major seafood commodities, including crustaceans, cephalopods and other mollusk species |
| Starchy Roots | Potatoes, Sweet Potatoes, Yams, Cassava, Taro, etc. |
| Fruit | Apples, bananas, oranges, berries, grapes, watermelon, pineapples, mangoes, peaches,<br>pears, cherries, kiwifruit, papayas, avocados, plums |
| Other meat | Game Meat (deer, elk, and moose), Exotic Meats (kangaroo, ostrich, and alligator), Rabbit,<br>Horse, Camel, Guinea Pig, Snails |

Hypertension prevalence (estimated share of the population with hypertension in adults aged 30-79) for 194 countries or regions from 1990-2019 (https://ourworldindata.org/grapher/hypertension-adults-30-79?time=latest), diabetes prevalence (the share of people aged 20-79 who have diabetes) for 190 countries or regions from 1980-2014 (https://ourworldindata.org/grapher/diabetes-prevalence-who-gho), and the estimated average level of non-high-density lipoprotein cholesterol (non-HDL-C, mmol/L) for 190 countries or regions from 1980-2018 (https://ourworldindata.org/grapher/average-non-hdl-cholesterol-levels) were obtained from Our World in Data (https://ourworldindata.org) database. Likewise, to ensure that all data were relatively complete while covering a sufficiently long period of time and corresponding to the dietary group data, hypertension prevalence for 159 countries or regions from 1992-2017, diabetes prevalence from 1992-2014, average non-HDL-C levels for 159 countries or regions from 1992-2017 were selected and averaged. Gross Domestic Product (GDP) data for these 159 countries or regions from 1992-2017 were similarly obtained and averaged (https://ourworldindata.org/grapher/gdp-per-capita-maddison-project-database).

### 2.3 Statistical analysis

#### 2.3.1 Correlation and partial correlation analysis

The Pearson correlation coefficients (n = 159) between each of 16 dietary components intake per capita (1992-2017 average) and hypertension prevalence (1992-2017 average), diabetes prevalence (1992-2014 average), and average non-HDL-C levels (1992-2017 average) were calculated by SPSS (V 25), respectively. Regression analyses of each dietary component with hypertension prevalence, diabetes prevalence, and average non-HDL-C levels were performed by Prism 10 respectively. GDP per capita was taken as a control variable, and partial correlation coefficients between intake of the 16 dietary components and hypertension prevalence, diabetes prevalence, and average non-HDL-C levels were calculated using SPSS (V 25).

#### 2.3.2 Temporal lag analysis

The associations between dietary components and health indicators showed consistent significant positive or negative correlations in both one star and two stars analyses were performed temporal association analysis (three stars analysis). Eighteen countries, including China, the United States of America, and Australia, were randomly selected as examples for the analyses. The temporal trajectories of each dietary component (1961-2020) with three health indicators (hypertension prevalence, 1990-2019; diabetes prevalence, 1980-2014; and average non-HDL-C levels, 1980-2018) were examined to assess their similarity of trends. In countries or regions exhibiting a consistent trend pattern, the presence of a stable time lag between trends in dietary components and trends in health indicators was judged. For each pair of analyses, a temporal association between the dietary component and the health indicator was considered to exist if there was a clear similarity in certain countries or regions and a relatively stable time lag was present.

### 2.4 Data visualization

Radar plots were plotted by Python to visualize the strength of one star and two stars associations between each of the 16 dietary components and hypertension prevalence, diabetes prevalence, and average non-HDL-C levels. Scatter plots with linear regression lines were plotted using Prism 10. In addition, temporal change area plots in the three stars analyses were plotted by Prism 10. Geographic heat maps of 159 countries or regions for hypertension prevalence (1992-2017 average), diabetes prevalence (1992-2014 average) and average non-HDL-C levels (1992-2017 average) were plotted with Tableau Desktop 2022.2. The graphical abstract and TSGMA flowcharts were created using bioGDP^24^ (https://biogdp.com), and the global MA networks framework diagram was created using BioRender (https://www.biorender.com).

## 3. Results

### 3.1 TSGMA of the associations between dietary components and average non-HDL-C levels

One star evaluation (Figure 2a, 2b) revealed that average non-HDL-C levels were significantly positively correlated with the intake of 11 dietary components, including red meat (Correlation Coefficient, CC = 0.566, *p* < 0.001), milk (CC = 0.596, *p* < 0.001), animal fats group (CC = 0.597, *p* < 0.001), sugar and sweeteners (CC = 0.676, *p* < 0.001), eggs (CC = 0.721, *p* < 0.001), fish and seafood (CC = 0.224, *p* < 0.001), nuts (CC = 0.316, *p* < 0.001), vegetable oils (CC = 0.438, *p* < 0.001), alcoholic beverages (CC = 0.448, *p* = 0.005), vegetables (CC = 0.452, *p* < 0.001), and poultry meat (CC = 0.492, *p* < 0.001). Conversely, significant negative correlations were observed for pulses (CC = −0.339, *p* < 0.001) and starchy roots (CC = −0.386, *p* < 0.001).

**Figure 2.**
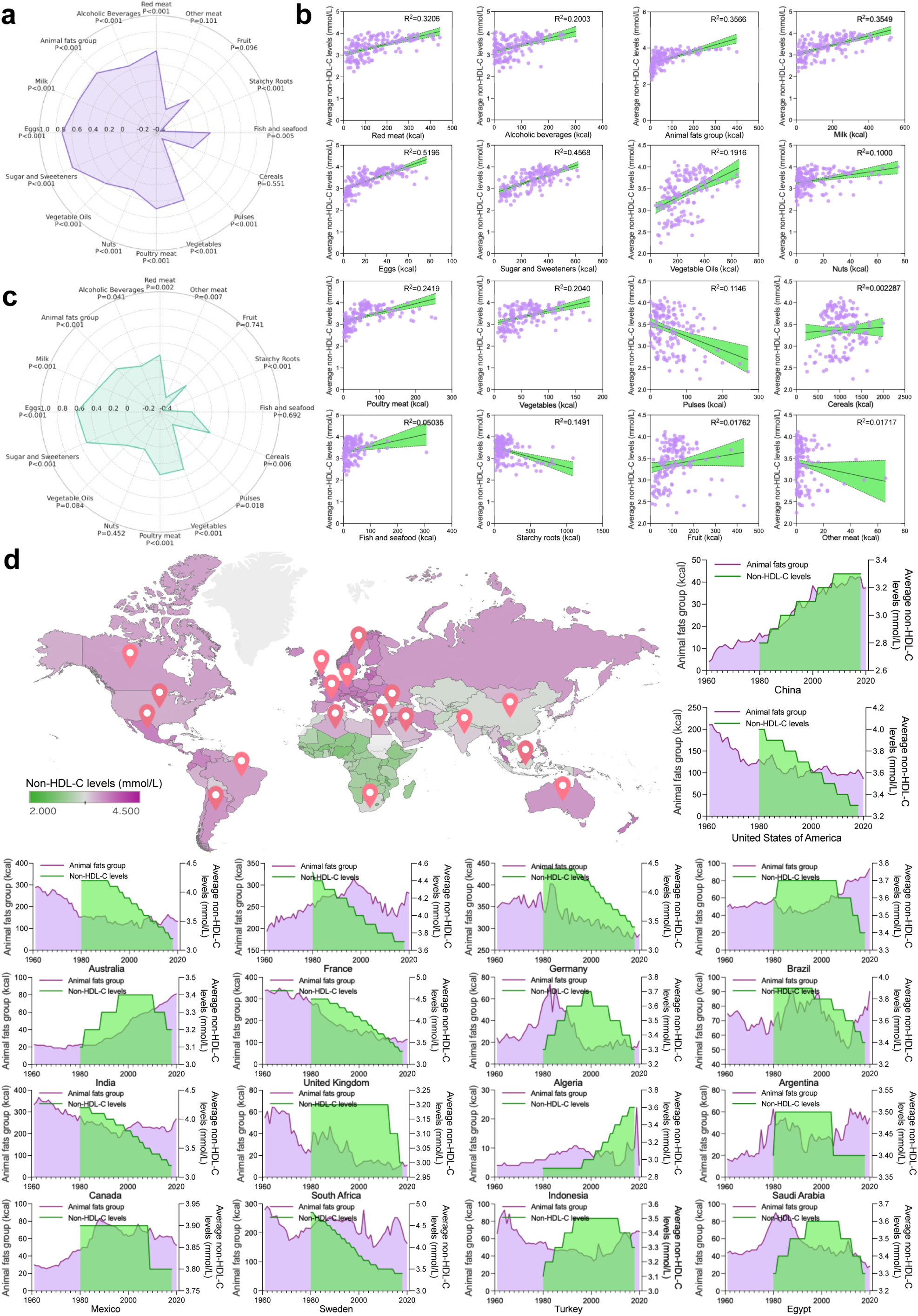
TSGMA analysis of associations between dietary components and average non-HDL-C levels. (a) Radar plot shows one star evaluation of Pearson correlation coefficients between the intake of 16 dietary components and average non-HDL-C levels across 159 countries or regions. (b) Scatter plot illustrates the association between dietary components and average non-HDL-C levels. (c) Radar plot shows two stars evaluation after adjusting for GDP per capita, based on partial correlation coefficients. (d) Three stars analysis of temporal trends. The geographic heatmap depicts the average non-HDL-C levels (1992-2017 averages) in 159 countries or regions worldwide, with particular highlights of the 18 countries randomly selected for the temporal analysis. The area plot shows temporal trends in animal fats group intake (purple) and average non-HDL-C levels (green) from 1961 to 2020.

Since GDP per capita is closely related to various life quality factors such as dietary structure, health care and education level, it can be considered as a representation of confounding factors in the global analysis^23^. Therefore, I choose the GDP per capita as an adjustment variable for the two stars analysis here. Following adjustment for GDP per capita (Figure 2c), average non-HDL-C levels remained significantly positively associated with 8 dietary components: alcoholic beverages (Partial Correlation Coefficients, PCC = 0.172, *p* = 0.041), animal fats group (PCC = 0.308, *p* < 0.001), vegetables (PCC = 0.317, *p* < 0.001), poultry meat (PCC = 0.328, *p* < 0.001), milk (PCC = 0.361, *p* < 0.001), sugar and sweeteners (PCC = 0.527, *p* < 0.001), eggs (PCC = 0.572, *p* < 0.001), and red meat (PCC = 0.259, *p* = 0.002). Negative associations with starchy roots (PCC = −0.361, *p* < 0.001) and pulses (PCC = −0.199, *p* = 0.018) also persisted after adjustment. Notably, associations with nuts (PCC = 0.064, *p* = 0.452), vegetable oils (PCC = 0.145, *p* = 0.084), and fish and seafood (PCC = −0.034, *p* = 0.692) became non-significant. Additionally, compared to the previous one star analysis, cereals (previously non-significant) showed a positive association (PCC = 0.231, *p* = 0.006), while other meats (previously non-significant) showed a significant negative association (PCC = −0.227, *p* = 0.007).

Animal fats group, red meat, eggs, alcoholic beverages, milk, poultry meat, pulses, starchy roots, sugar and sweeteners and vegetables, which were already significantly associated with both one star and two stars analysis, were selected for the three stars analysis (as described in Section 2.2.1).

For animal fats group, among the 18 randomly selected countries, China, the United States, Australia, Germany, the United Kingdom, Algeria, Canada, and Egypt showed similar trends between animal fats group intake and non-HDL-C levels, and changes in non-HDL-C levels lagged behind animal fats group intake changes (Figure 2d). For instance, in China, both animal fats group intake and non-HDL-C levels have continued to rise; in the United States, both have declined. In Algeria, animal fat group intake peaked around 1982 and began to decline thereafter, with non-HDL-C levels also peaking and declining about 16 years later. Similarly, in Egypt, animal fats group intake peaked around 1980 and non-HDL-C levels peaked around 1994 and began to decline after 2005. In summary, in countries or regions where changes in animal fats group intake follow a similar trend to that of non-HDL-C levels, there is always a lag of about 15 years.

For red meat, similar lagged relationships were observed in China, the United States, Australia, the United Kingdom, Canada, Argentina, and Indonesia (Figure S1). For example, in Australia, red meat intake began declining around 1972, with non-HDL-C levels following a downward trend approximately 19 years later. In the United Kingdom, red meat intake declined from around 1971, with non-HDL-C levels decreasing about 16 years later. In Germany, a decline in red meat intake starting in 1987 preceded a fall in non-HDL-C levels by approximately six years. In Indonesia, a rise in red meat intake around 1978 was followed by an increase in non-HDL-C levels around 1996. Overall, the lag between red meat intake changes and non-HDL-C levels response ranged from 15 to 20 years.

For eggs, similar lagged patterns were observed in the United States, Australia, the United Kingdom, Germany, and Canada (Figure S2). In Australia, a decline in egg consumption around 1978 was followed by a decrease in non-HDL-C levels around 1991 (lag approximately 13 years). In Germany, egg intake declined around 1980, with non-HDL-C levels falling around 1993 (lag approximately 13 years). Collectively, the lag between egg intake and non-HDL-C levels changes consistently approximated 13 years.

By contrast, trends in alcoholic beverages (Figure S3), milk (Figure S4), poultry meat (Figure S5), pulses (Figure S6), starchy roots (Figure S7), sugar and sweeteners (Figure S8), and vegetables (Figure S9) were not similar to those for non-HDL-C levels in the majority of countries or regions.

In summary, based on TSGMA, nuts, vegetable oils, and fish and seafood are one star risk factors for non-HDL-C levels. Alcoholic beverages, vegetables, poultry meat, milk, sugar and sweeteners are two stars risk factors for non-HDL-C levels, while starchy roots and pulses are two stars protective factors for non-HDL-C levels. Animal fats group, red meat, and eggs are three stars risk factors for non-HDL-C levels. Moreover, cereals, fruit and other meat are zero star risk factors, showing no association with non-HDL-C levels.

### 3.2 TSGMA of the associations between dietary components and diabetes prevalence

One star evaluation (Figure 3a, 3b) showed that diabetes prevalence was significantly positively correlated with the intake of three dietary components: poultry meat (CC = 0.326, *p* < 0.001), cereals (CC = 0.287, *p* < 0.001), and sugar and sweeteners (CC = 0.228, *p* = 0.004). In contrast, significant negative correlations were observed for alcoholic beverages (CC = −0.329, *p* < 0.001) and starchy roots (CC = −0.238, *p* = 0.003).

**Figure 3.**
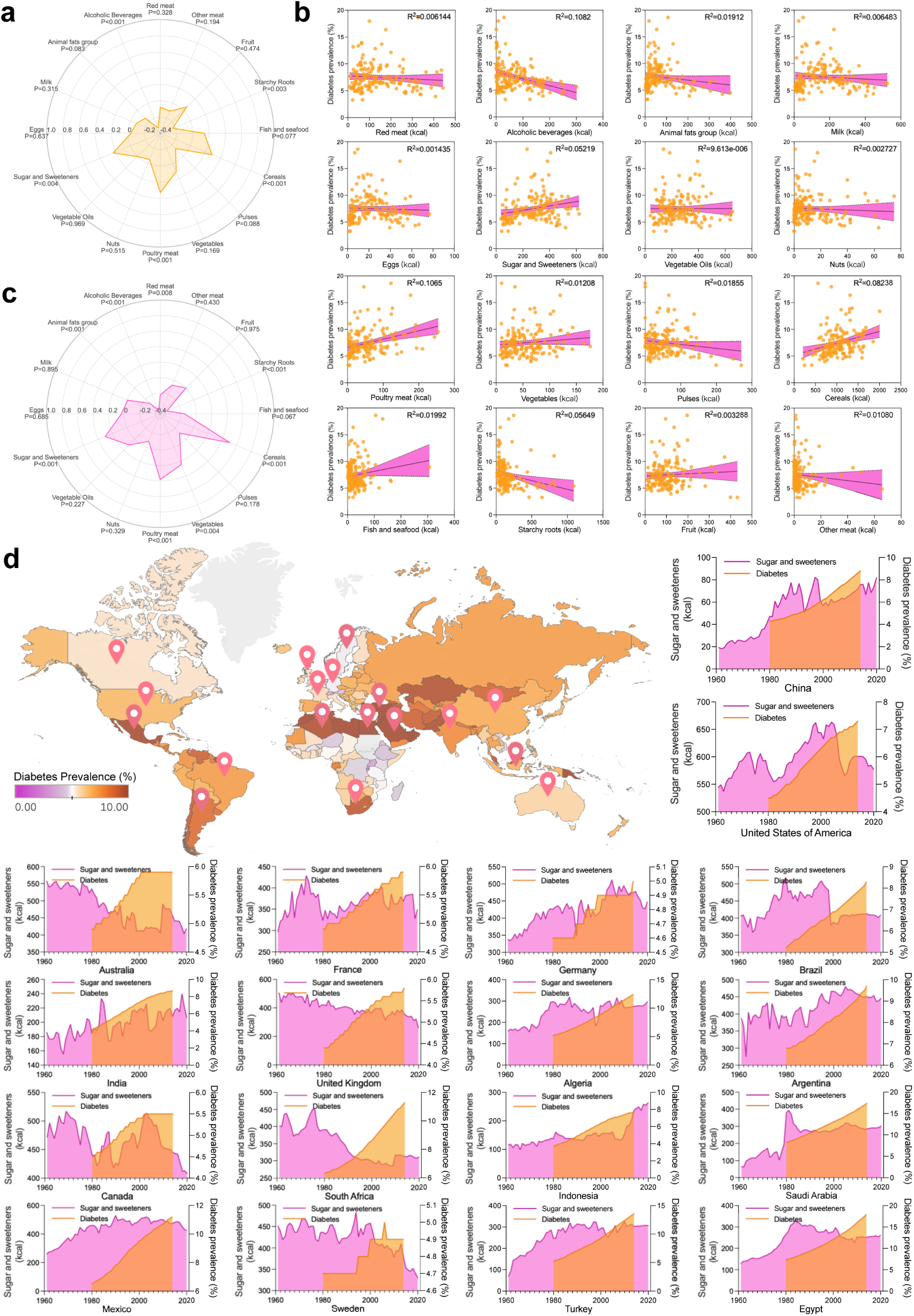
TSGMA analysis of associations between dietary components and diabetes prevalence. (a) Radar plot shows one star evaluation of Pearson correlation coefficients between the intake of 16 dietary components and diabetes prevalence across 159 countries or regions. (b) Scatter plot illustrates the association between dietary components and diabetes prevalence. (c) Radar plot shows two stars evaluation after adjusting for GDP per capita, based on partial correlation coefficients. (d) Three stars analysis of temporal trends. The geographic heatmap shows the diabetes prevalence (1992-2014 averages) in 159 countries or regions worldwide, with particular highlights of the 18 countries randomly selected for the temporal analysis. The area plot shows temporal trends in sugar and sweeteners intake (pink) and diabetes prevalence (orange) from 1961 to 2020.

After adjustment for GDP per capita (Figure 3c), positive associations between diabetes prevalence and cereals (PCC = 0.504, *p* < 0.001), poultry meat (PCC = 0.380, *p* < 0.001), and sugar and sweeteners (PCC = 0.301, *p* < 0.001) remained significant. Negative associations with alcoholic beverages (PCC = −0.419, *p* < 0.001) and starchy roots (PCC = −0.374, *p* < 0.001) also persisted. Notably, vegetables, which were non-significant in one star analysis, became positively associated with diabetes after adjustment (PCC = 0.243, *p* = 0.004). In addition, animal fats group (PCC = –0.275, *p* = 0.001) and red meat (PCC = −0.223, *p* = 0.008) emerged as significant negative associations only after adjustment. In both the one star and two stars analysis, sugar and sweeteners, cereals, poultry meat, starchy roots, and alcoholic beverages were all consistently significantly associated with the diabetes prevalence, and they were selected for the three stars analysis. For sugar and sweeteners (Figure 3d), seven countries, including China, Germany, Brazil, Saudi Arabia, Mexico, Turkey, and Egypt, exhibited parallel upward trends in sugar and sweetener consumption and diabetes prevalence, with diabetes prevalence changes lagging behind diabetes prevalence intake trends. For cereals, ten countries, including China, the United States, India, South Africa, Algeria, Mexico, Indonesia, Saudi Arabia, and Egypt, demonstrated similar patterns: rising cereal intake was followed by increases in diabetes prevalence (Figure S10).

Similarly, for poultry meat, the majority of the 18 randomly selected countries showed consistent upward trends in poultry meat intake and subsequent rises in diabetes prevalence, indicating a high level of temporal coherence (Figure S11). In contrast, for starchy roots, China, France, Germany, Brazil, Argentina, and Sweden displayed opposing trends between intake and diabetes prevalence, with increased consumption associated with lower diabetes prevalence (Figure S12). However, for alcoholic beverages, no consistent temporal pattern linked to diabetes prevalence was observed across the majority of countries or regions (Figure S13). Moreover, the specific value of the time lag in the three stars analysis of the relationship between diabetes prevalence and dietary components is difficult to determine because the vast majority of these countries or regions have no obvious transitions or features that can be used to quantitatively determine the lag time.

Thus, in the relationship between dietary components and diabetes, poultry meat, cereals, and sugar and sweeteners are three stars risk factors for diabetes, while starchy roots are three stars protective factors. Two stars association factor include only alcoholic beverages with protective effects, no dietary components are found as one star risk factors for diabetes. Moreover, red meat, animal fats group, milk, eggs, vegetable oils, nuts, vegetables, pulses, fish and seafood, fruit and other meat are zero star risk factors, showing no association with diabetes prevalence.

### 3.3 TSGMA of the associations between dietary components and Hypertension

One star evaluation (Figure 4a, 4b) showed that hypertension prevalence was significantly positively associated with milk intake (CC = 0.256, *p* = 0.001) and alcoholic beverage intake (CC = 0.220, *p* = 0.006), while a significant negative correlation was observed with fish and seafood consumption (CC = −0.194, *p* = 0.015).

**Figure 4.**
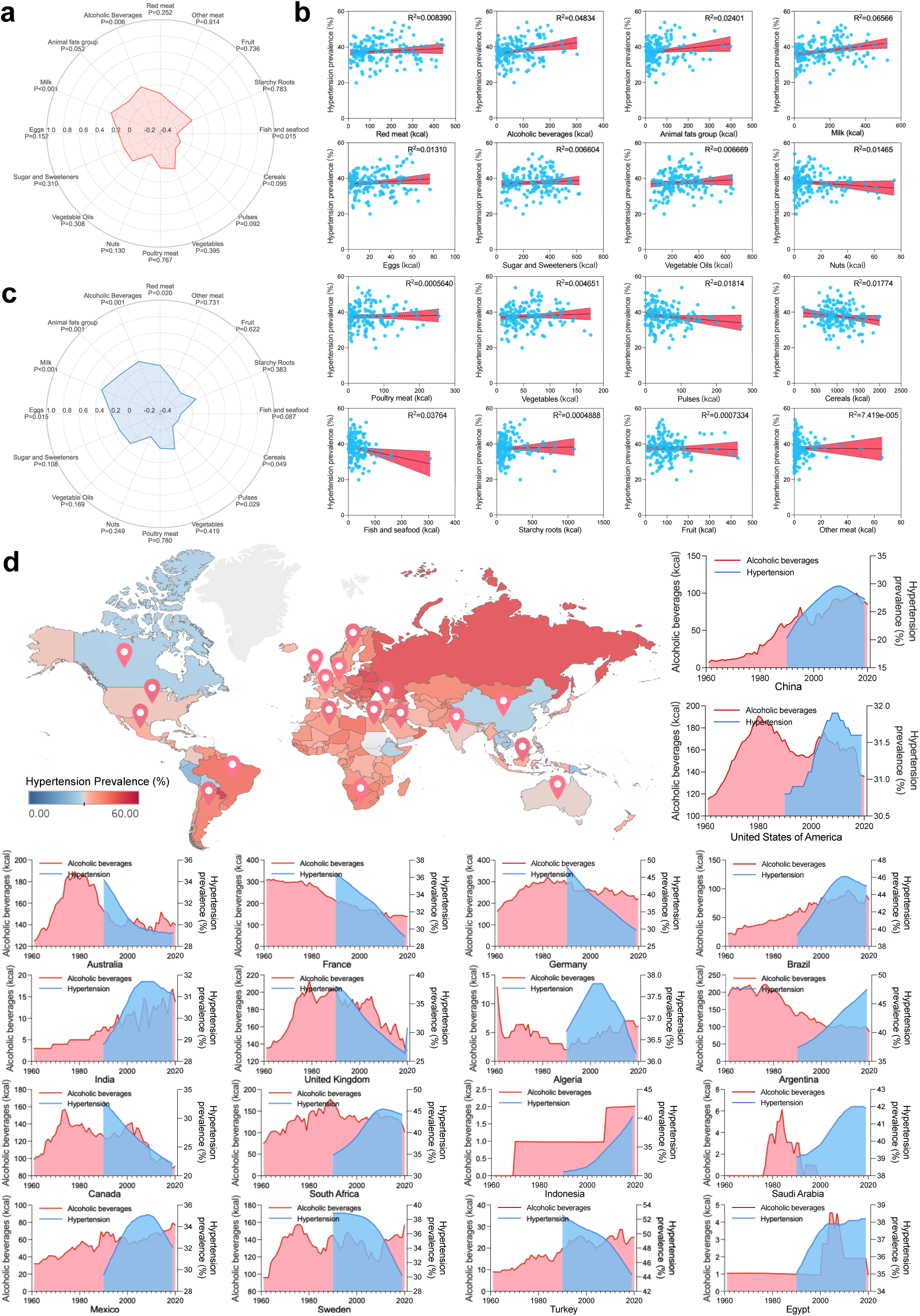
TSGMA analysis of associations between dietary components and hypertension prevalence. (a) Radar plot shows one star evaluation of Pearson correlation coefficients between the intake of 16 dietary components and average hypertension prevalence across 159 countries. (b) Scatter plot illustrates the association between dietary components and hypertension prevalence. (c) Radar plot showing two stars evaluation after adjusting for GDP per capita, based on partial correlation coefficients. (d) Three stars analysis of temporal trends. The geographic heatmap shows the hypertension prevalence (1992-2017 averages) in 159 countries worldwide, with particular highlights of the 18 countries randomly selected for the temporal analysis. The area plot shows temporal trends in alcoholic beverages intake (red) and hypertension prevalence (blue) from 1961 to 2020.

Following adjustment for GDP per capita, two stars analysis (Figure 4c) revealed that hypertension prevalence remained significantly positively associated with milk (PCC = 0.379, *p* < 0.001) and alcoholic beverages (PCC = 0.296, *p* < 0.001). However, the previously observed negative correlation with fish and seafood intake was no longer significant (PCC = −0.144, *p* = 0.087).

Additionally, several dietary components not associated at the one star analysis became significant after adjustment: animal fats group (PCC = 0.288, *p* = 0.001), eggs (PCC = 0.203, *p* = 0.015), and red meat (PCC = 0.195, *p* = 0.020) were positively correlated with hypertension prevalence, while cereals (PCC = −0.166, *p* = 0.049) and pulses (PCC = −0.184, *p* = 0.029) showed significant negative associations.

In the TSGMA of dietary components and hypertension prevalence, only alcoholic beverages and milk were consecutively significantly associated with hypertension prevalence in one star and two stars analyses, thus alcoholic beverages and milk were selected for three stars analysis. For alcoholic beverages (Figure 4d), eight countries, including China, the United States, Australia, France, Brazil, India, Canada, and South Africa, exhibited similar temporal trends between alcoholic beverages intake and hypertension prevalence. In most cases, increases in alcoholic beverage consumption preceded subsequent rises in hypertension prevalence. For example, in China, alcoholic beverage intake rose steadily from around 1980, while hypertension prevalence showed a corresponding upward trend since before around 2010. In the United States, alcoholic beverages intake continued to rise, with a turnaround in 1980, while the turnaround in the increase of hypertension prevalence occurred around 2008, with a time lag of about 28 years. In Australia, alcoholic beverages intake showed a turnaround from declining to holding steady in 1996, while hypertension prevalence showed a similar turnaround around 2010, with a lag of about 16 years. In France, a long-term decline in alcohol consumption paralleled a decline in hypertension prevalence over the same period. In South Africa, the turnaround in the rise in alcoholic beverages intake occurred around 1984, and a similar turnaround in hypertension prevalence occurred around 2010, with a lag of approximately 26 years. By contrast, for milk, most countries or regions did not display a coherent temporal relationship between milk intake and hypertension prevalence (Figure S14). Together, these findings suggest that alcoholic beverage consumption may contribute to changes in hypertension prevalence with a temporal lag, whereas no consistent time-lagged relationship was observed for milk intake.

Therefore, alcoholic beverages are three stars risk factors for hypertension, milk is a two stars risk factor, while Fish and seafood are found as a one star protective factor. However, red meat, animal fats group, eggs, sugar and sweeteners, vegetable oils, nuts, poultry meat, vegetables, pulses, cereals, starchy roots, fruit and other meat are zero star risk factors, showing no association with hypertension prevalence.

## 4. Discussion

### 4.1 Macro associations

Comprehensive analysis of global-scale associations within extensive datasets is essential for understanding risk factors linked to human health^20,25,26^. The rapid expansion of data availability and computational capabilities has fundamentally altered how data are collected, processed, and utilized^27–29^. Traditional concepts of "big data" may no longer fully capture the scale, depth, or reliance upon contemporary datasets, which continue to expand exponentially.

Here, I introduce the term “macro data” (MD) to characterize datasets that are vast in scale, multidimensional in structure, and broadly cross. In the context of MD, the association based on these data also rises to a completely new dimension, and decoding all the associations behind these data will fundamentally change our understanding of the world. Therefore, I name the association of all data behind MD as “macro association” (MA), and I name the exploration of MA as “macro association analysis” (MA-analysis). The essence of MA-analysis is to rely on MD to systematically integrate maximum possible relevant variables to reveal comprehensive multidimensional associations. Given the unprecedented volume, granularity and analytical potential of current data, MA-analysis will provide a deeper, previously unattainable understanding of the world.

Extensive adoption of MA-analysis can lead to the construction of global MA networks. MA-analysis can operate at various scales, from micro-level associations such as molecular or cellular networks (comprehensive gene or protein interaction networks) to macro-level relationships involving global phenomena, including global dietary, climate, economic, or health^30–35^. Moreover, cross-level analyses integrating micro and macro scales through suitable MA methodologies offer additional layers of insight into complex global dynamics.

Among multiple potential applications, global MA-analysis stands out due to its direct relevance to human life, health, economics, and societal dynamics. The macro-level data is particularly advantageous when compared to the intricacies of molecular-level data because of the relative simplicity of the calculations associated with processing predominantly numerical and easily interpretable data^36,37^. Furthermore, databases such as Our World in Data (https://ourworldindata.org) and growing public accessibility to global datasets have now created optimal conditions for implementing global MA-analysis.

In epidemiological research, conventional correlation or regression analyses, despite their immediacy, often yield inconclusive or misleading results due to confounding factors^38–42^. Advanced epidemiological methodologies, including case-control studies, cohort studies, and meta-analyses, while more reliable and valid, are resource-intensive and time-consuming^42–44^, and most are not suitable for analyzing global data on a country-by-country basis. As a result, these rigorous methods typically focus on confirming established or controversial relationships for which the data are suitable, whereas extensive and rapid exploration of potential associations is not and equally unsuitable for MA-analysis.

However, rapid, efficient and accurate analysis is essential for MA-analysis. A large amount of country-level global data is currently underutilized, despite being readily available from various sources. Therefore, there is an urgent need for streamlined, efficient, and relatively accurate methods to bridge the gap between immediate but superficial association analyses and rigorous but resource-intensive epidemiological studies, as well as to conduct and systematize MA-analysis in preparation for the establishment of global MA networks.

### 4.2 Three Stars Global Macro Association-analysis

Given the exponential growth in global data availability, there is an urgent need for a simple, rapid, and reliable analytical approach to facilitate MA-analysis. Here, I propose the Three Stars Global Macro Association-analysis (TSGMA), a simple but accurate method designed to efficiently classify associations between risk factors through a straightforward three-step evaluation process. TSGMA categorizes associations using progressively rigorous analytical steps: correlation analysis (one star), partial correlation analysis controlling for confounders (two stars), and temporal relationship assessment for potential causality (three stars).

The one star analysis initially identifies potential associations through correlation coefficients calculated from averaged national-level data across multiple years. Although straightforward, correlation analysis swiftly highlights possible association on a global scale. However, correlations alone cannot distinguish genuine associations from spurious ones influenced by confounding variables such as socioeconomic conditions, policy changes, or technological advancements^23,45–47^. Thus, further refinement is essential.

For the two stars analysis, critical confounders that could significantly bias the correlation are identified and controlled via partial correlation analysis. Given that overly extensive adjustments can inadvertently mask true associations, it is crucial to select only the most significant and representative confounders to maintain accuracy^48,49^. Moreover, broad one star analysis can also assist in the finding of critical confounder. In brief, Variables consistently exhibiting two stars associations indicate robust relationships worthy of further consideration and investigation.

In the final three stars analysis, temporal relationships between risk factors and health outcomes are assessed at the national level. Specifically, trends over time are analyzed to determine if consistent lagged relationships exist. Such temporal analyses provide stronger evidence for potential causality and further mitigate the impact of residual or unidentified confounders, enhancing the reliability of identified associations.

This comprehensive and systematic method significantly advances our ability to quickly and reliably evaluate associations from extensive global datasets. TSGMA has extensive applicability, ranging from resolving longstanding debates to uncovering novel associations across diverse research fields. Meanwhile, the objects analyzed by TSGMA can be transformed according to the data conditions, either in country or regional units, and are applicable to issues with similar data conditions. The multiple levels also allow variables with different degrees of association to be revealed, rather than a single criterion causing the omission of numerous weak or moderate associations. Moreover, a harmonized and standardized approach to quantifying these associations also makes them comparable. As an example, the TSGMA system I conducted in this study revealed associations between global dietary patterns and three major cardiometabolic health indicators (Non-HDL-C levels, diabetes prevalence, and hypertension prevalence). Among numerous dietary factors, animal fat, red meat, and eggs emerged as robust three stars risk factors strongly associated with elevated non-HDL-C levels and with lag times ranging from approximately 13 to 20 years. Notably, these findings are highly consistent with established evidence in nutritional epidemiology^50–52^. In contrast, pulses and starchy roots steadily demonstrated protective two stars associations, reflecting their role in lipid regulation due to their high fiber content and lower glycemic impact^53,54^. In the context of diabetes, poultry meat, cereals, and sugar and sweeteners exhibited strong, persistent three stars associations, aligning well with existing evidence highlighting refined carbohydrates and excessive poultry intake as potential diabetes risk factors through mechanisms involving insulin resistance and impaired glucose homeostasis^55–57^. Conversely, Starchy roots demonstrated a protective two stars association, as did with non-HDL-C levels^58^. In terms of hypertension prevalence, intake of alcoholic beverages showed a three stars risk association with hypertension. This strong association highlights the physiological pathways by which alcohol elevates blood pressure through mechanisms such as vasoconstriction and sympathetic activation^59,60^. Milk intake presented a two stars association, but lacked consistent temporal trends across most analyzed countries or regions, suggesting differential regional consumption patterns^61^. Interestingly, fish and seafood intake were identified as a one stars protective factor, aligning with prior literature that suggests a clear cardiovascular protective effect of omega-3 fatty acids from fish oil^62,63^. However, the association did not reach a higher level of robustness may due to the infrequent, geographically-specific, and idiosyncratic nature of fish and seafood consumption, as has been observed in previous studies indicating the instability of this association^62,64,65^. Moreover, I and Qi successfully applied a similar approach to systematically reassess the long-debated relationship between red meat consumption and cancer incidence previously^23^, providing robust conclusions from a novel analytical perspective.

Taken together, these findings underscore TSGMA’s utility in rapidly identifying dietary risk factors at a global scale, confirming known associations, and uncovering novel patterns that traditional epidemiological studies might miss due to methodological constraints. These results provide actionable insights for public health interventions targeting dietary modifications, highlighting areas needing further mechanistic exploration to substantiate causality and inform global dietary guidelines. Critically, TSGMA lays the foundation for future MA-analyses and the construction of comprehensive global MA networks by providing accurate assessments of association strength, clearly defining different levels of risk, and dramatically reducing computational and resource demands without sacrificing analytical rigor or accuracy.

### 4.3 Construction of global MA networks

TSGMA can be effectively employed using existing data frameworks organized by either specific research domains or databases. In the case of databases, currently, most global databases primarily function as repositories for data collection rather than platforms for integrated analytical insight^66–68^.

By introducing MA-analysis into public databases such as Our World in Data (https://ourworldindata.org), FAO (https://www.fao.org/faostat/en/#data/FBS), International Agency for Research on Cancer (https://www.iarc.who.int), and the World Bank (https://data.worldbank.org) or self-constructed datasets, we can systematically interconnect collected variables through TSGMA. Subsequent analyses utilizing artificial intelligence and advanced visualization techniques can facilitate the construction of intra-database MA networks. This transformative approach moves beyond traditional data aggregation towards integrated data analysis and visualization, representing a significant evolution in database functionality^66–68^.

As individual databases progressively establish their internal MA networks, inter-database integration can occur, linking networks across different data repositories. This interconnection allows for cross-validation of overlapping data and addresses gaps through complementary datasets, ultimately culminating in a unified global MA network. Moreover, these interconnected MA networks can serve as the foundation for developing a dedicated global MA networks database. Such a resource would systematically document and continuously update one star, two stars, and three stars associations among global data variables, profoundly enhancing our understanding of global dynamics and facilitating solutions to complex challenges such as climate change, disease epidemiology, economic fluctuations, and environmental shifts.

Each analytical level within TSGMA (from one to three stars) holds an important value. Even initial one star assessments, identifying potential correlations without confirmed causality, can offer significant insights into global patterns. This has important insights for the initial phase of MA-network construction. For example, constructing a one star MA network using Our World in Data (https://ourworldindata.org) could rapidly enhance our understanding of global relationships. Progressively incorporating two to three stars analyses and considering graded correlation strengths will further enrich these networks.

Looking ahead, a global MA-analysis initiative could systematically apply MA-analysis across all fields. I propose a structured framework for constructing MA networks (Figure 5): starting from TSGMA within individual databases or domains, progressing through inter-database or inter-domain networks, and culminating in comprehensive global MA networks. Ultimately, by integrating macro-level data (such as global socio-economic indicators) with micro-level biological data (such as proteins, nucleic acids), we can establish fully integrated domain-wide MA networks. This approach promises unprecedented insights into the interconnected dynamics shaping our world.

**Figure 5.**
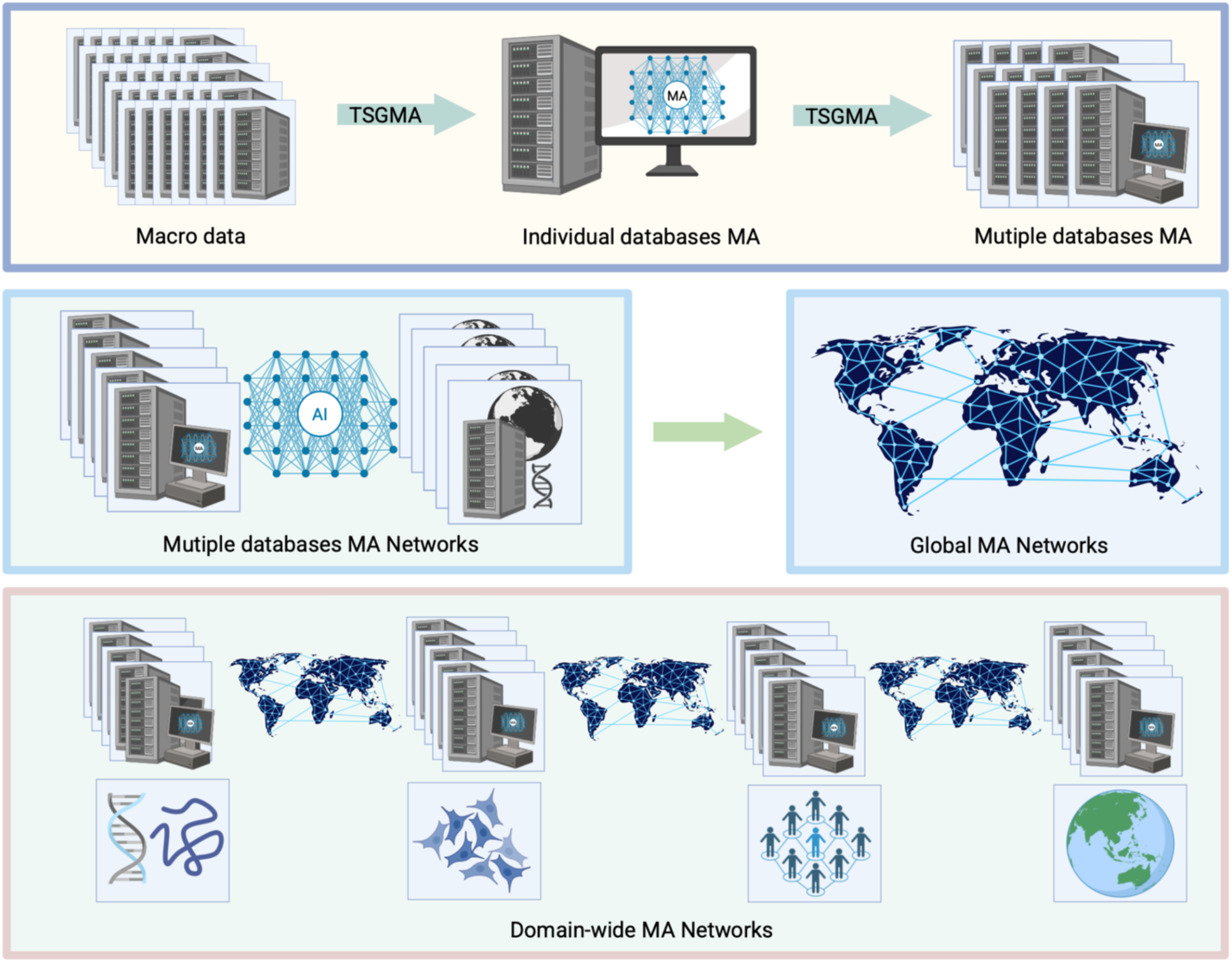
Global MA networks development framework

## 5. Conclusion

In this study, I introduce the concept of MA for the first time and highlight the urgency and importance of conducting MA-analysis in the context of rapidly expanding MD. To address this need, I developed the TSGMA, a novel framework designed to identify MA within increasingly available harmonized global datasets. TSGMA integrates correlation analysis, confounder adjustment, and temporal validation to efficiently and accurately classify associations into a standardized, graded system. When applied to global dietary and health indicator data, TSGMA successfully identified both well-established and previously overlooked associations, demonstrating its capability to extract actionable insights from complex global data.

Beyond its analytical function, TSGMA also provides a structural foundation for building MA networks. Leveraging both TSGMA and MA-analysis, I propose a framework for constructing global MA networks: starting from intra-database networks and progressing toward the integration of inter-database associations. This framework offers a unified platform for tracking, comparing, and interpreting global drivers of health, economy, and environment, etc.

Together, the integration of MA, TSGMA, and global MA networks establishes a structured and scalable approach to uncovering meaningful patterns from MD, providing a practical foundation for systematic discovery and hypothesis generation in global research.

### Limitations of the study

Despite its advantages, TSGMA has several limitations. First, precise adjustment for confounders remains challenging, requiring more systematic methods such as multivariate regression or principal component analysis. Second, variables initially non-significant in one star analysis but becoming significant after adjusting for confounders may provide overlooked insights. Third, temporal validation (three stars analysis) needs refinement; integrating machine learning could enhance accuracy in detecting time-lag relationships.

Moreover, to facilitate rapid and broad screening, I assessed associations between individual factors and outcomes without considering their interactions. Although major confounders were controlled, potential inaccuracies from correlated factors remain; for instance, dietary patterns often link meat intake with alcohol consumption, an interaction were also observed in this study. Future analyses should further incorporate such interactive or cumulative effects. Finally, integrating automated artificial intelligence approaches with TSGMA is urgently needed to further enhance the efficiency of MA-analysis.

## Data Availability

All data produced are available online at [Database: https://ourworldindata.org]

## RESOURCE AVAILABILITY

### Lead contact

Further information and reasonable requests for resources and reagents should be directed to and will be fulfilled by the lead contact, Hongyue Ma.

### Materials availability

This study did not generate new samples or unique reagents.

#### Data and code availability

***• Data***: This paper analyzes existing, publicly available data, accessible at [Database: https://ourworldindata.org].
***• Code***: This paper does not report original code.
***• Additional Information***: Any additional information required to reanalyze the data reported in this article is available from the lead contact upon request.

## ACKNOWLEDGMENTS

This work is mainly supported by Hongyue Ma’s personal funds and resources.

## AUTHOR CONTRIBUTIONS

Hongyue Ma performed all analyses and wrote the manuscript.

