## Supplemental Figure for "TSGMA: identification of macro associations from global data to build global MA networks"

Hongyue Ma[1, 2, 3]

1. Haide College, Ocean University of China, Qingdao, China.

2. School of Life Sciences, Westlake University, Hangzhou, Zhejiang, China.

3. Lead contact

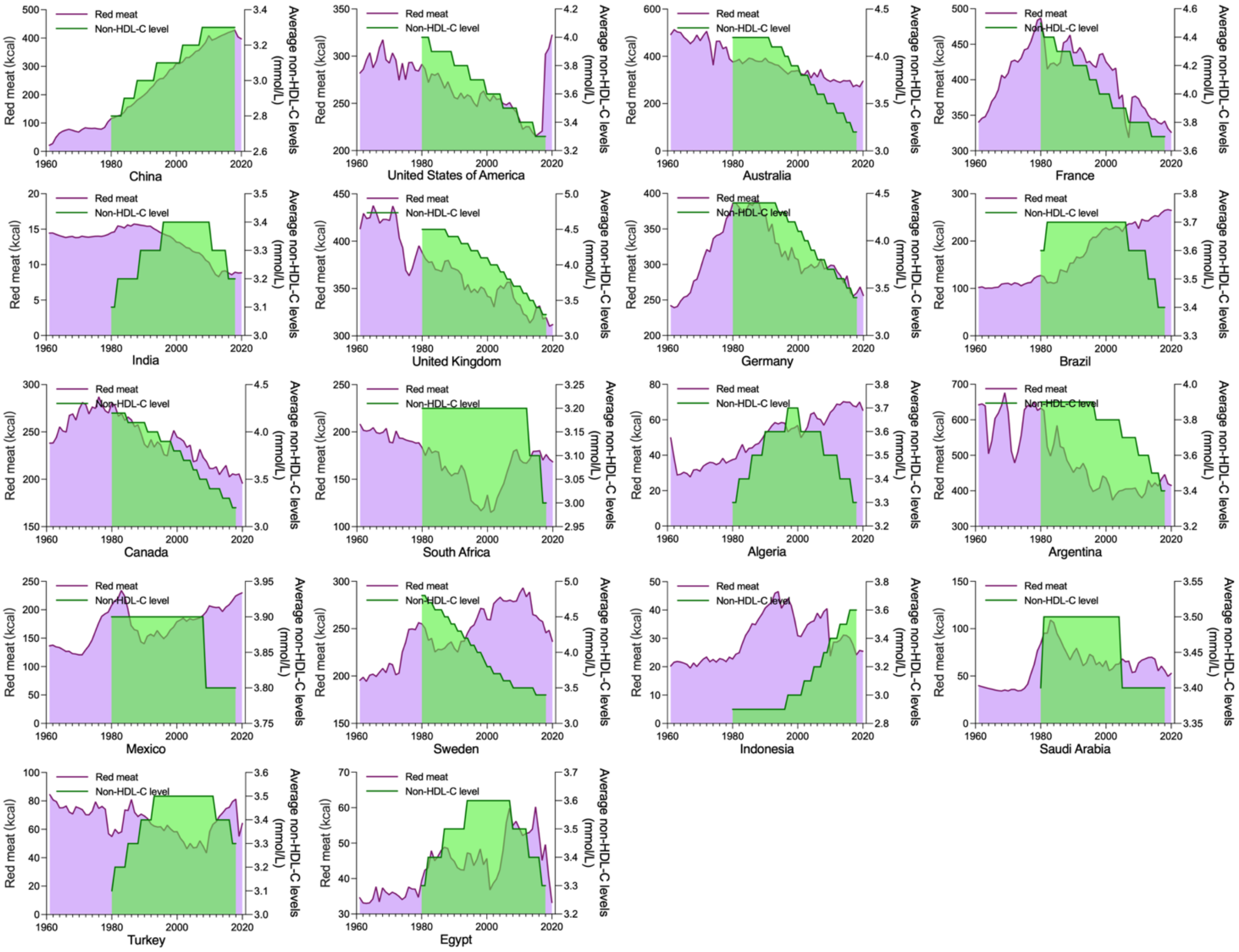


**Figure S1.** The temporal changes in red meat intake and non-HDL-C levels.

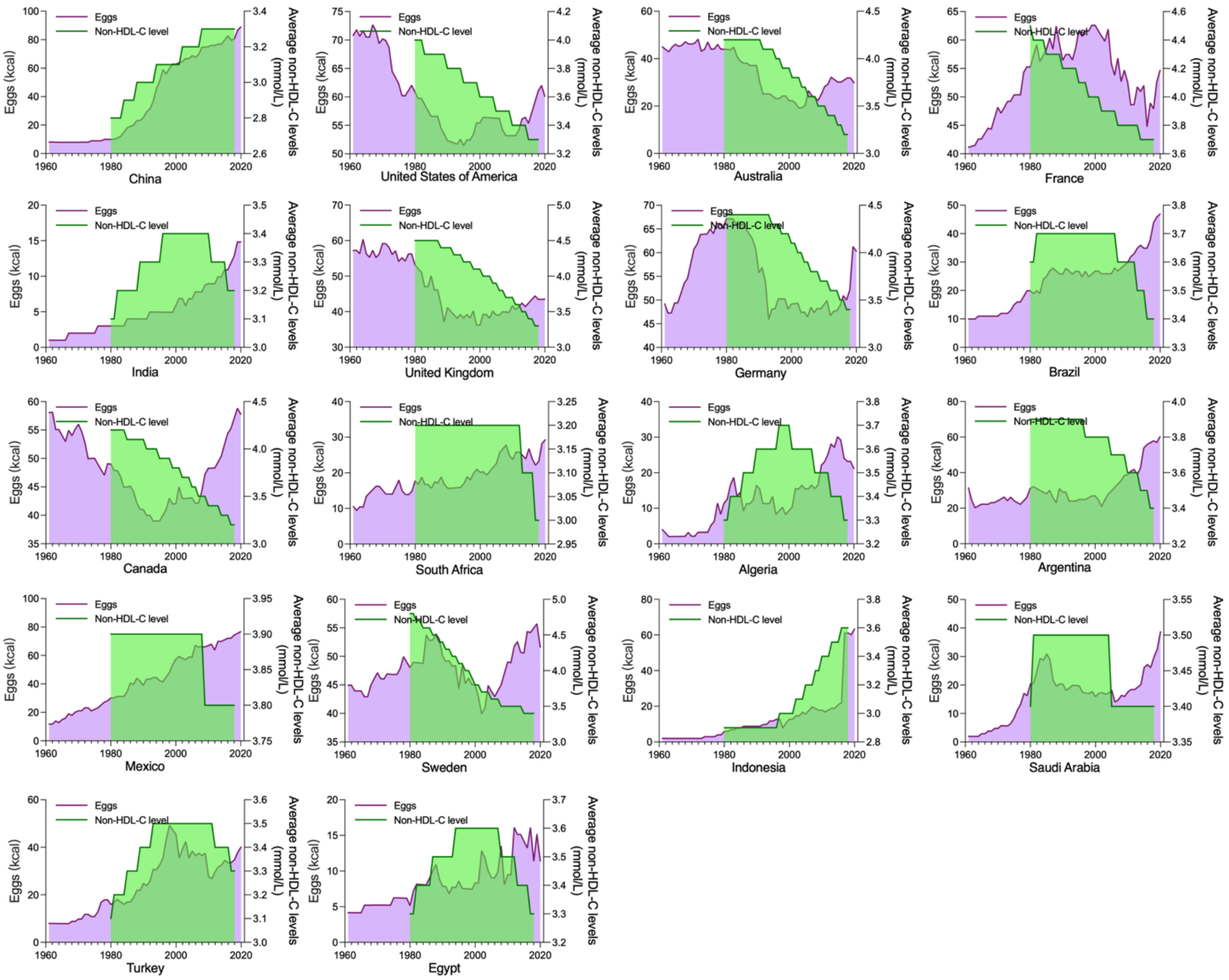


**Figure S2.** The temporal changes in eggs intake and non-HDL-C levels.

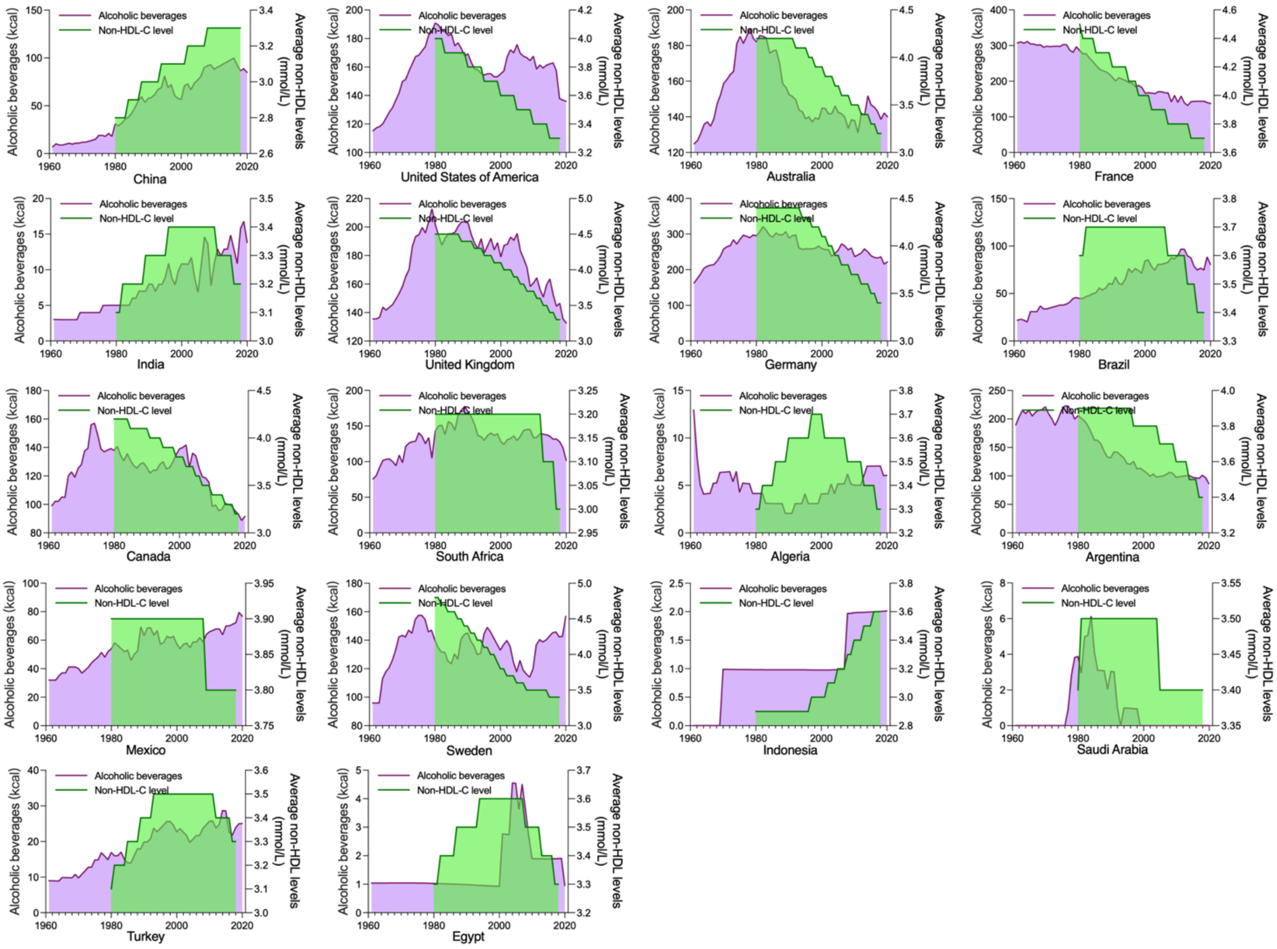


**Figure S3.** The temporal changes in alcoholic beverages intake and non-HDL-C levels.

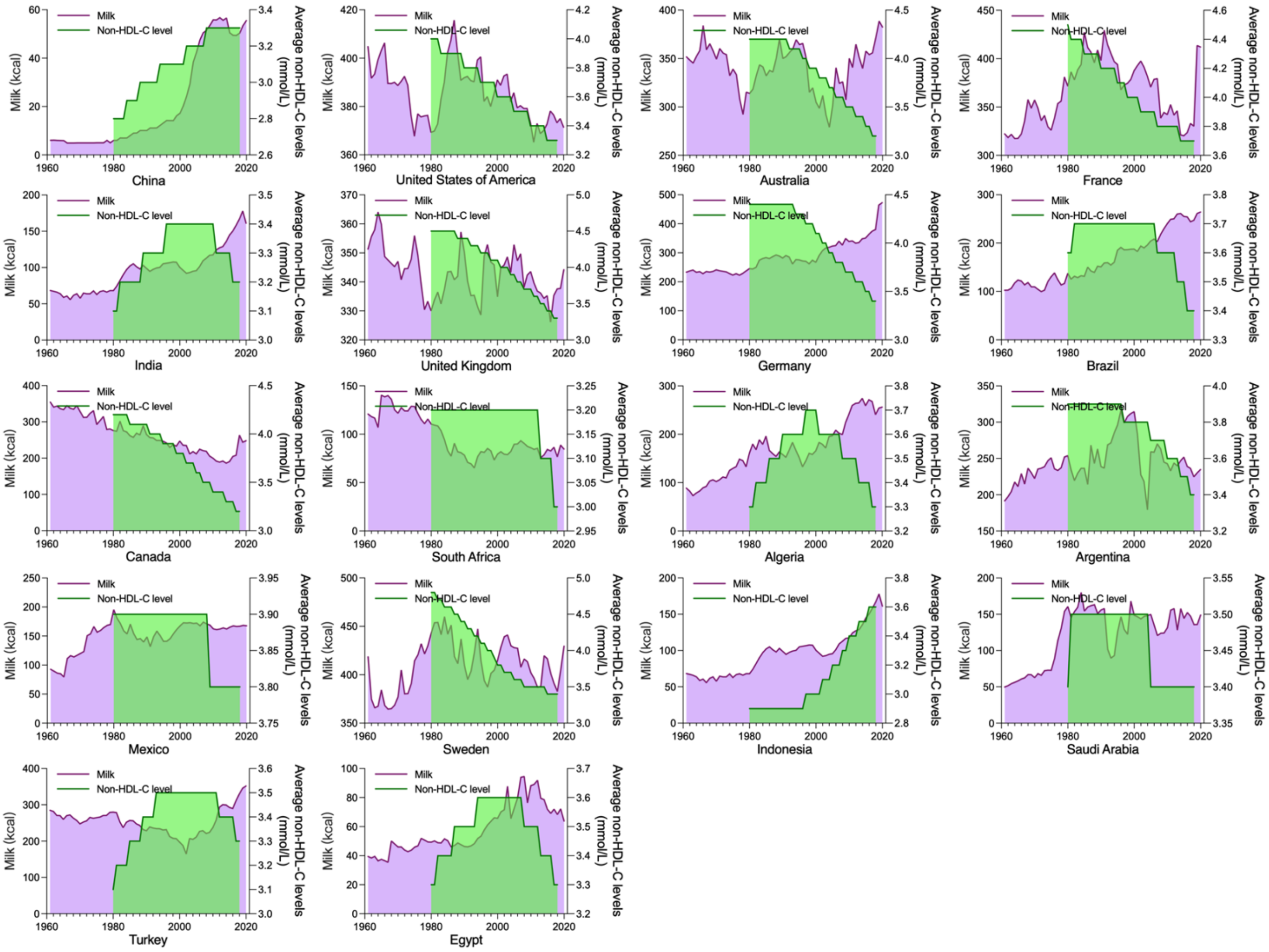


**Figure S4.** The temporal changes in milk intake and non-HDL-C levels.

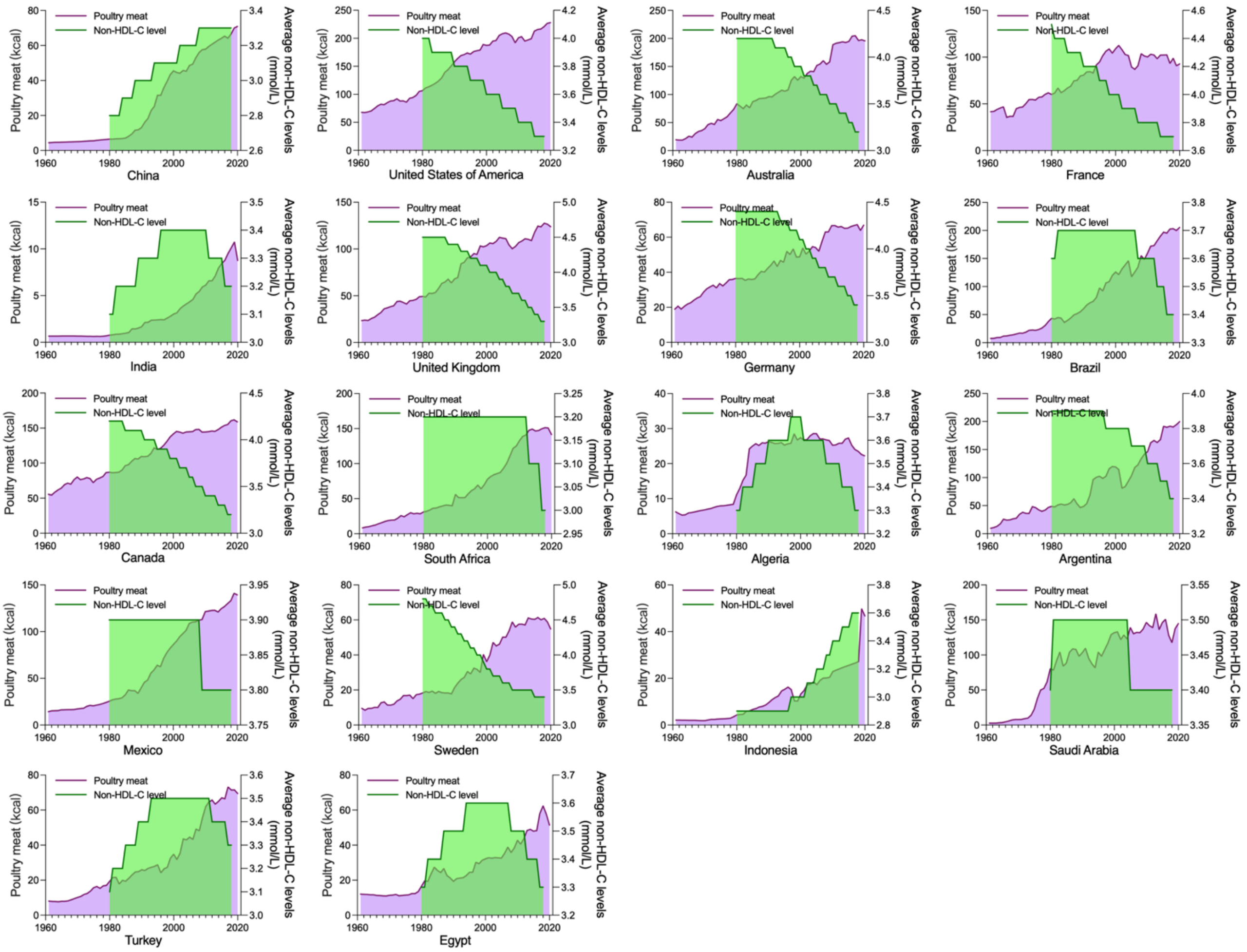


**Figure S5.** The temporal changes in poultry meat intake and non-HDL-C levels.

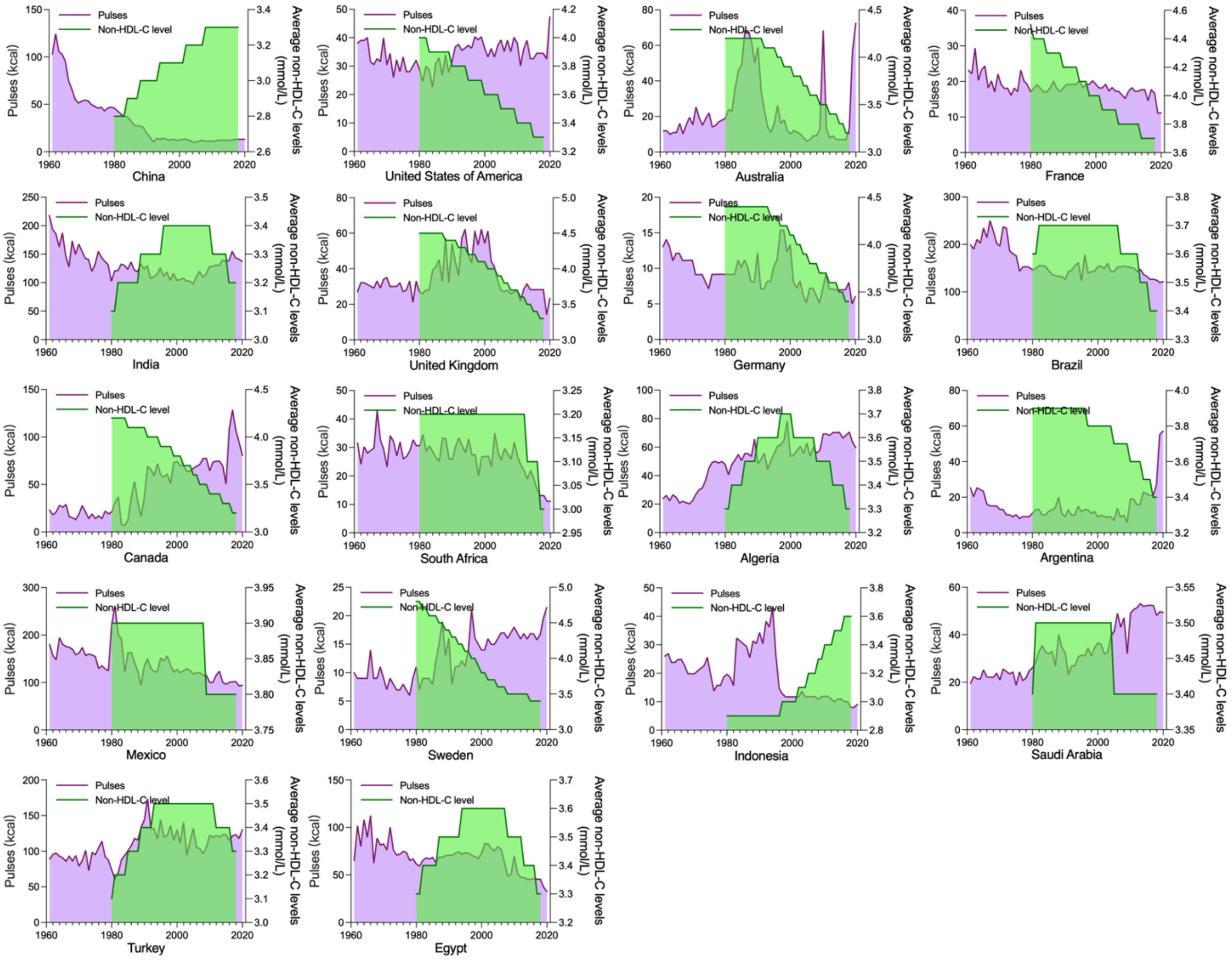


**Figure S6.** The temporal changes in pulses intake and non-HDL-C levels.

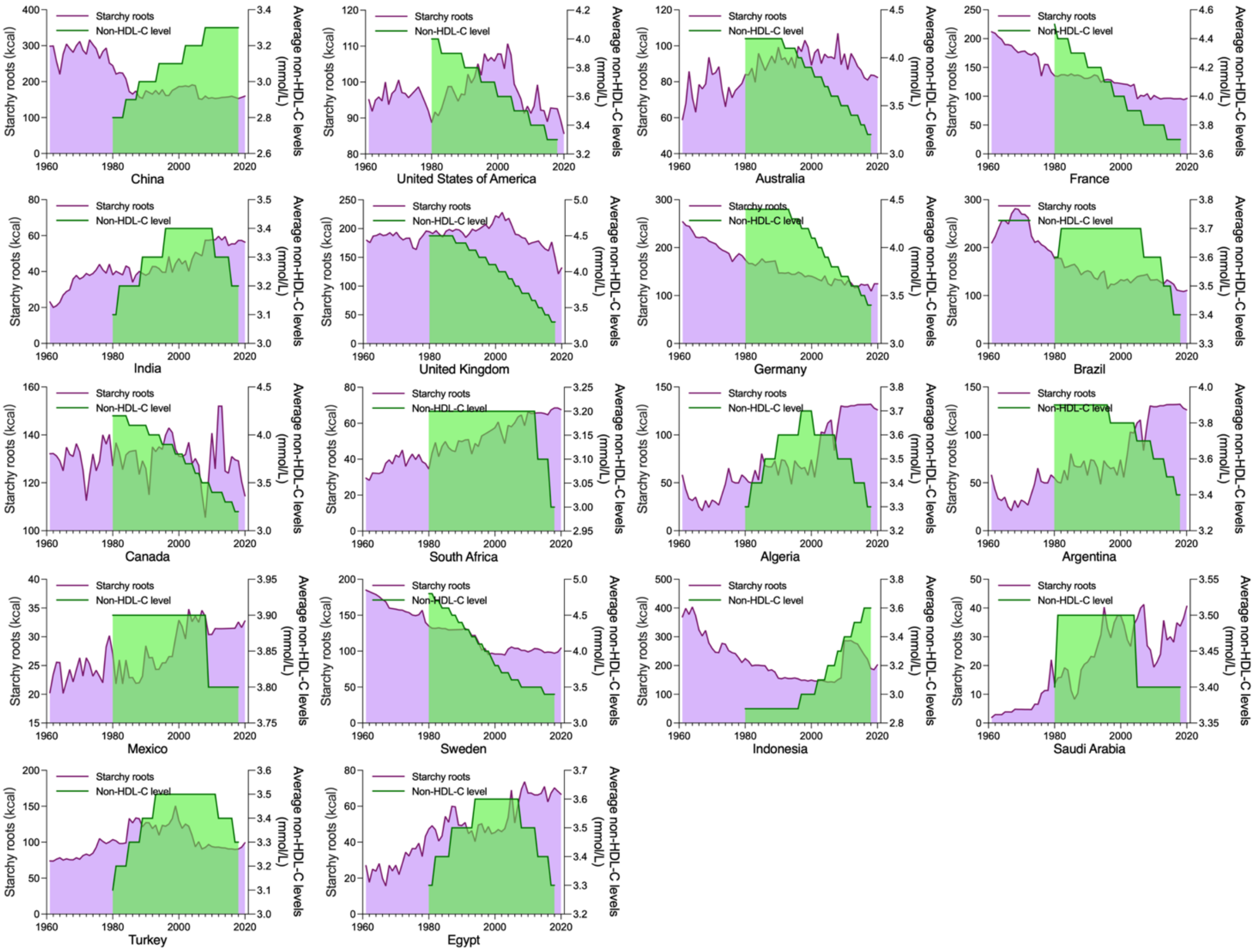


**Figure S7.** The temporal changes in starchy roots intake and non-HDL-C levels.

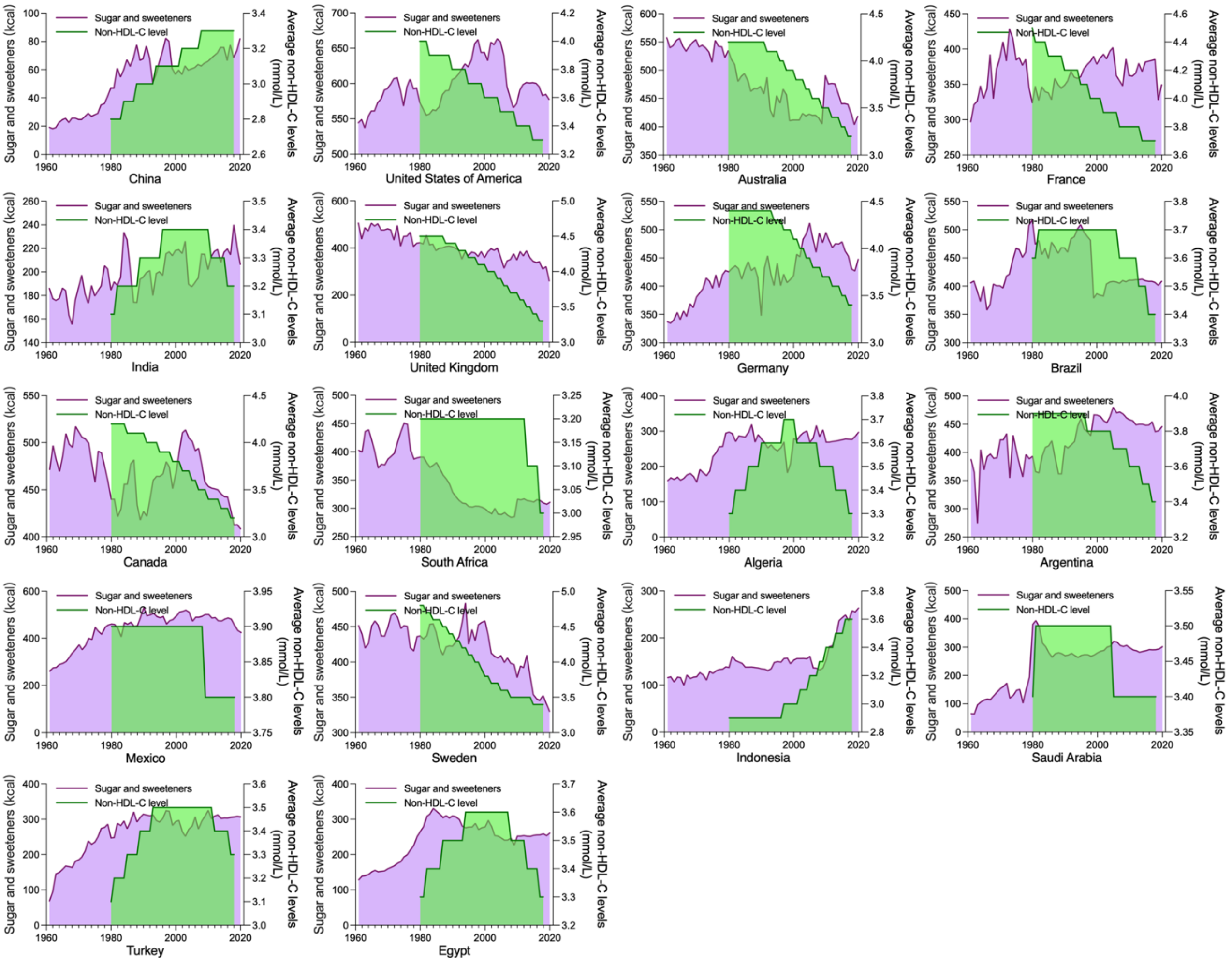


**Figure S8.** The temporal changes in sugar and sweeteners intake and non-HDL-C levels.

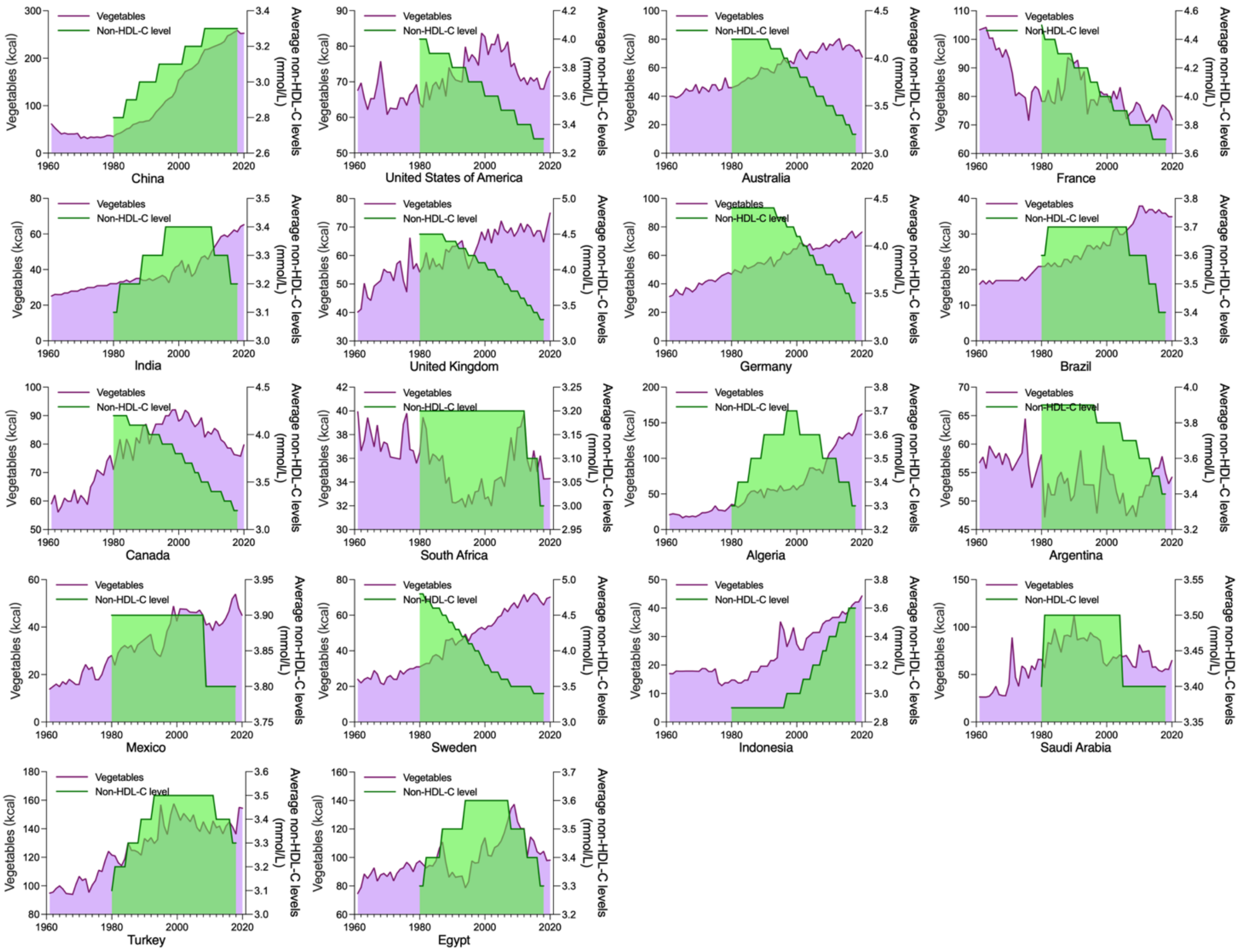


**Figure S9.** The temporal changes in vegetables intake and non-HDL-C levels.

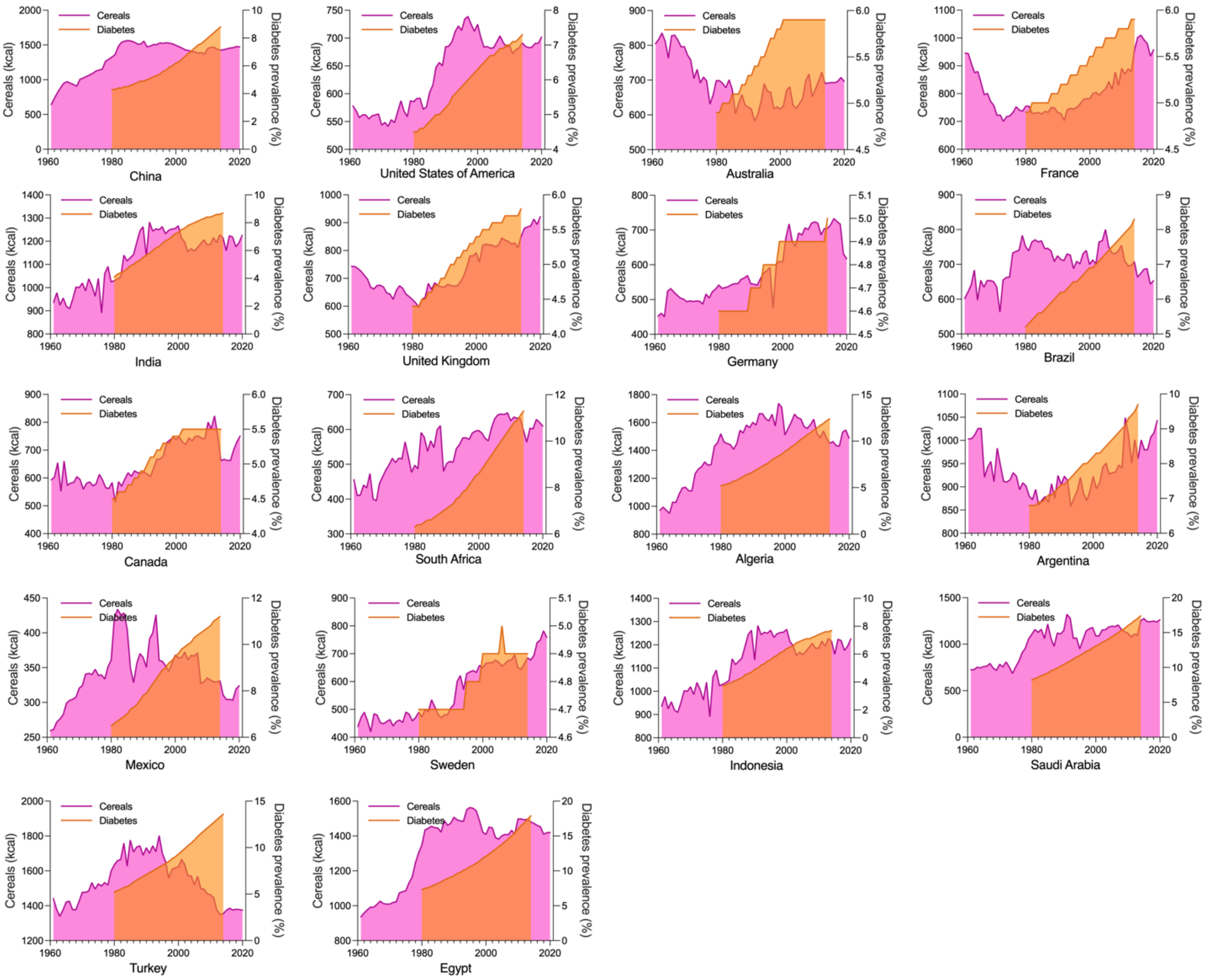


**Figure S10.** The temporal changes in cereals intake and diabetes prevalence.

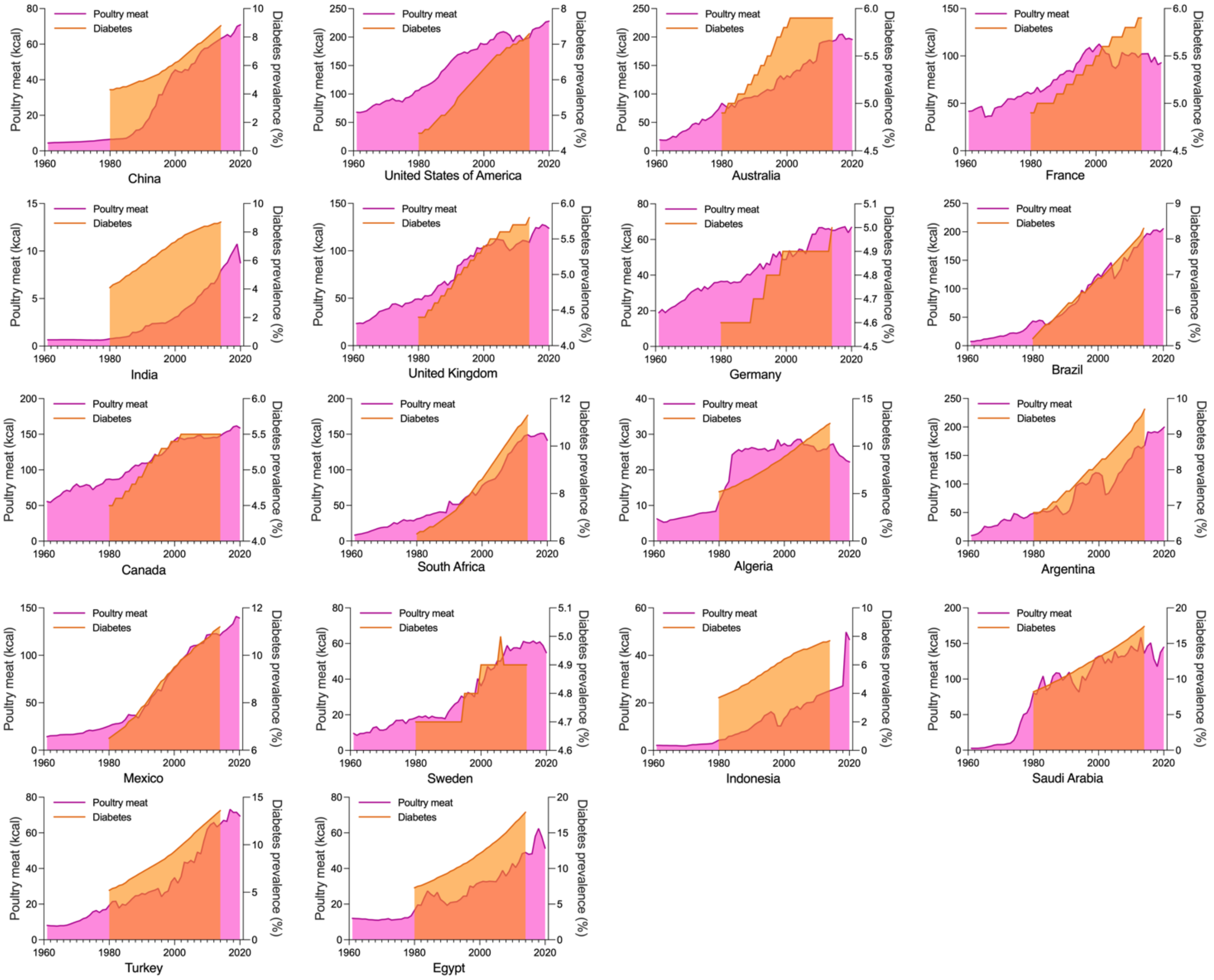


**Figure S11.** The temporal changes in poultry meat intake and diabetes prevalence.

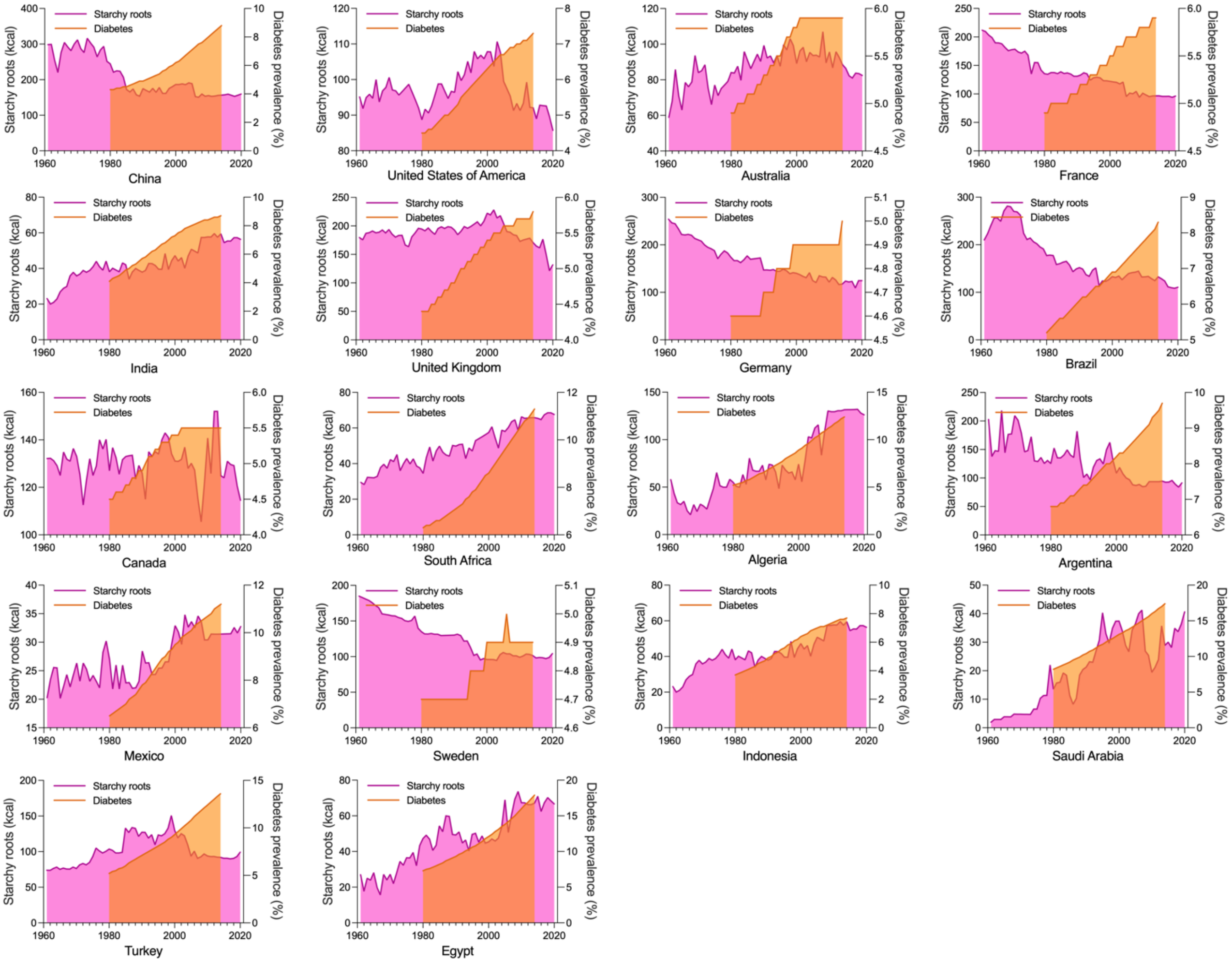


**Figure S12.** The temporal changes in starchy roots intake and diabetes prevalence.

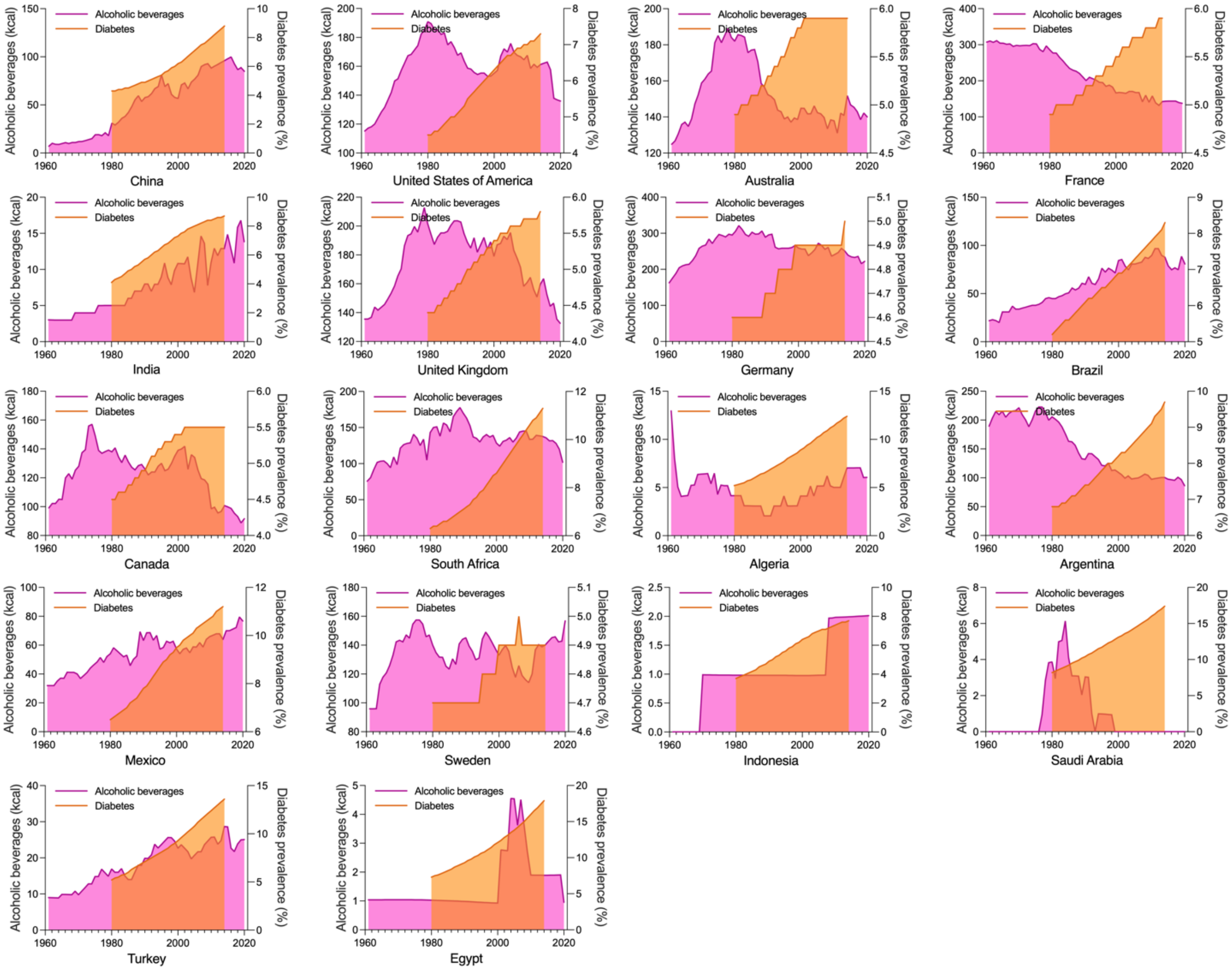


**Figure S13.** The temporal changes in alcoholic beverages intake and diabetes prevalence.

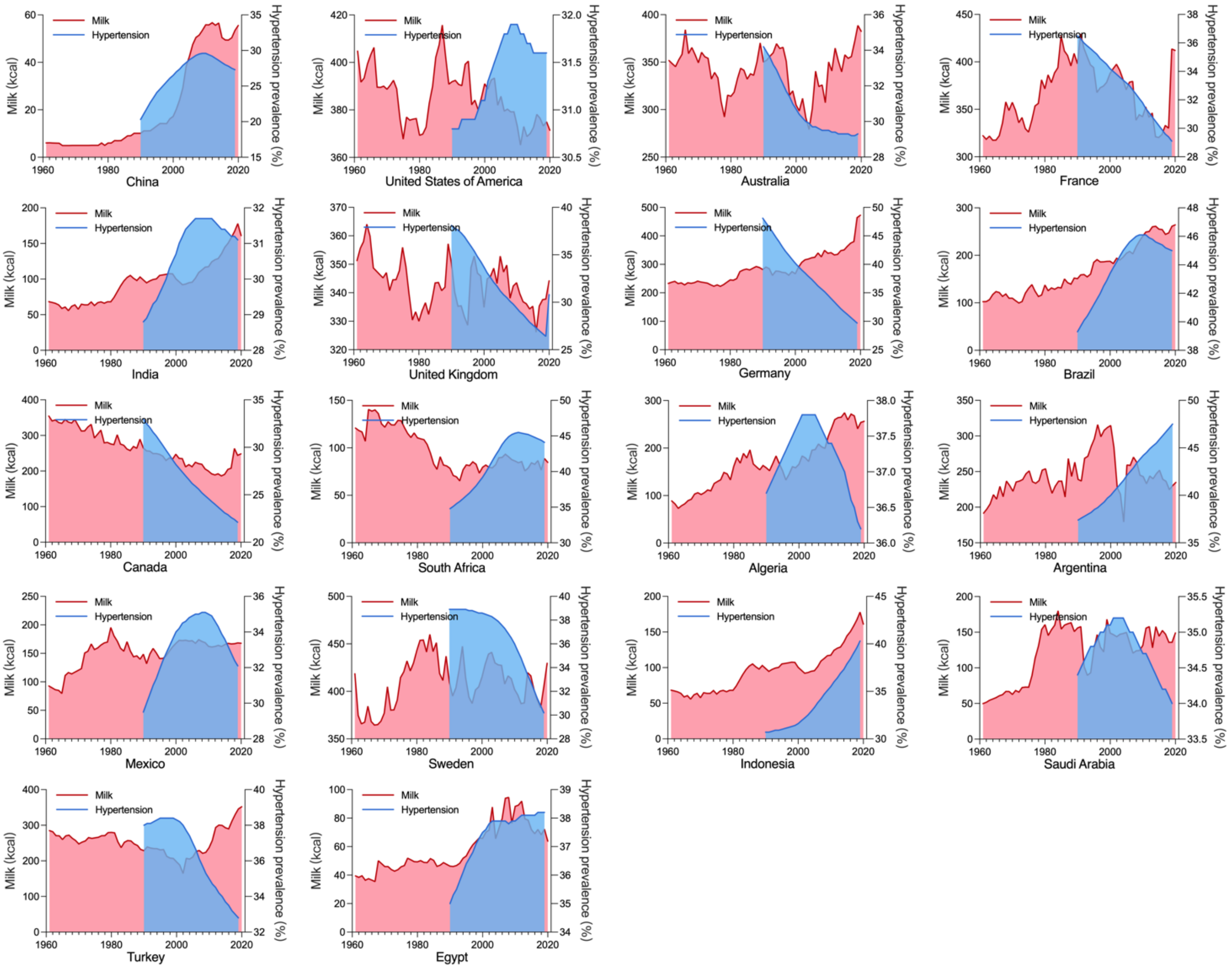


**Figure S14.** The temporal changes in milk intake and hypertension prevalence.
